# Dynamics and ecology of a multi-stage expansion of Oropouche virus in Brazil

**DOI:** 10.1101/2024.10.29.24316328

**Authors:** Houriiyah Tegally, Simon Dellicour, Jenicca Poongavanan, Carla Mavian, Graeme Dor, Vagner Fonseca, Massimiliano S. Tagliamonte, Marcel Dunaiski, Monika Moir, Eduan Wilkinson, Carlos Frederico Campelo de Albuquerque, Livia C. V. Frutuoso, CLIMADE Consortium, Edward C. Holmes, Cheryl Baxter, Richard Lessells, Moritz U.G. Kraemer, José Lourenço, Luiz Carlos Junior Alcantara, Tulio de Oliveira, Marta Giovanetti

## Abstract

In March 2024, the Pan American Health Organization (PAHO) issued an alert in response to a rapid increase in Oropouche fever cases across South America. Brazil has been particularly affected, reporting a novel reassortant lineage of the Oropouche virus (OROV) and expansion to previously non-endemic areas beyond the Amazon Basin. Utilising phylogeographic approaches, we reveal a multi-scale expansion process with both short and long-distance dispersal events, and diffusion velocities in line with human-mediated jumps. We identify forest cover, banana and cocoa cultivation, temperature, and human population density as key environmental factors associated with OROV range expansion. Using ecological niche modelling, we show that OROV circulated in areas of enhanced ecological suitability immediately preceding its explosive epidemic expansion in the Amazon. This likely resulted from the virus being introduced into simultaneously densely populated and environmentally favourable regions in the Amazon, such as Manaus, leading to an amplified epidemic and spread beyond the Amazon. Our study provides valuable insights into the dispersal and ecological dynamics of OROV, highlighting the role of human mobility in colonisation of new areas, and raising concern over high viral suitability along the Brazilian coast.

## Main text

Oropouche virus (OROV; *Oropouche orthobunyavirus*) is an arthropod-borne virus first identified in 1955 in Oropouche, a village in Trinidad and Tobago (*1*). OROV typically causes a febrile illness with symptoms such as high fever, headache, myalgia, arthralgia, photophobia, nausea, vomiting, and dizziness (*2*). In some cases, the illness can progress to severe neurological complications, including meningo-encephalitis (*1*). This re-emerging virus circulates primarily among wildlife such as non-human primates, rodents, sloths, and birds. It has caused around 30 documented human outbreaks in the Amazon region in recent years (*3*, *4*). The midge *Culicoides paraensis* serves as the primary vector for human transmission, but other secondary vectors include *Culex quinquefasciatus, Coquillettidia venezuelensis,* and *Aedes (Ochlerotatus) serratus (1, 5)*.

In March 2024, the Pan American Health Organization (PAHO) issued an alert in response to a rapid increase in Oropouche fever cases across several countries, including Brazil, Cuba, Bolivia, Colombia, and Peru (*6*, *7*). By October 6, 2024, a total of 10,275 confirmed cases of Oropouche had been reported across nine countries in the Americas, as well as the first two deaths (*6*, *8*). Brazil has been particularly affected, reporting not only the highest number of cases, but also severe complications linked to Oropouche virus infection (*9*). Recent epidemiological data and genomic investigations in Brazil (*10*, *11*) have described the recent expansion of OROV into previously non-endemic regions. These studies have identified reassortment events in the virus genome that may have contributed to its changing epidemiology. While the exact role of reassortment in the adaptation of OROV to novel environments remains to be fully understood, it has possibly impacted its spread into new ecological niches (*10*, *11*).

As with other arboviruses (*12*), recent changes in ecological context, such as deforestation, urbanisation, human mobility, and climate change, have possibly contributed to the emergence of OROV in new regions (*13*). In particular, environmental disruption pushes non-human mammal reservoirs and vectors into closer contact with human populations, facilitating viral spread (*14*). Additionally, human activities (*15*, *16*) like urban expansion and altered land use increase the risk of transmission of OROV in peri-urban and urban settings, where vectors such as *Culicoides paraensis* and *Culex quinquefasciatus* thrive (*17*–*19*). Despite these insights, significant gaps remain in quantifying the precise impact of these factors on recent OROV transmission dynamics. In addition, much remains unknown about the broader disease ecology of OROV, particularly concerning environmental correlates of local circulation and of its recent expansion. This lack of comprehensive understanding hampers effective risk assessment and preparedness efforts, both within Brazil and across the Americas.

This study aims to formally test key epidemiological hypotheses regarding OROV disease ecology and its range expansion. We integrate spatially explicit pathogen genomes and epidemiological data with geospatial data in a phylodynamic and ecological niche modelling framework to (i) reconstruct the dispersal history of OROV lineages across Brazil and analyse dispersal statistics in the context of a range expansion, (ii) evaluate the environmental factors associated with OROV transmission during distinct transmission phases, and (iii) map the ecological niche of OROV transmission to identify covariates of circulation suitability in the context of the expansion, and to pinpoint surveillance blind spots.

## Results

### Dispersal history and dynamics of OROV in Brazil

The epidemiological dynamics of OROV expansion in Brazil in late 2023 and 2024 show a two-stage process: a rapid rise in cases in Amazonian states, particularly Manaus, followed by widespread circulation in other parts of the country (Supplementary Figure S1). To further investigate, we applied a continuous phylogeographic approach using over ∼500 genomes sampled between 2022 and 2024 (*10*, *11*). Building on prior knowledge of reassortment events across genome segments (*10*, *11*), we conducted separate phylogeographic reconstructions for segments L, M, and S. Our analysis extracted spatiotemporal data from 100 annotated trees subsampled from post burn-in posterior distributions, revealing new insights into transition events between sampled regions (**Figure 1A**).

**Figure 1.**
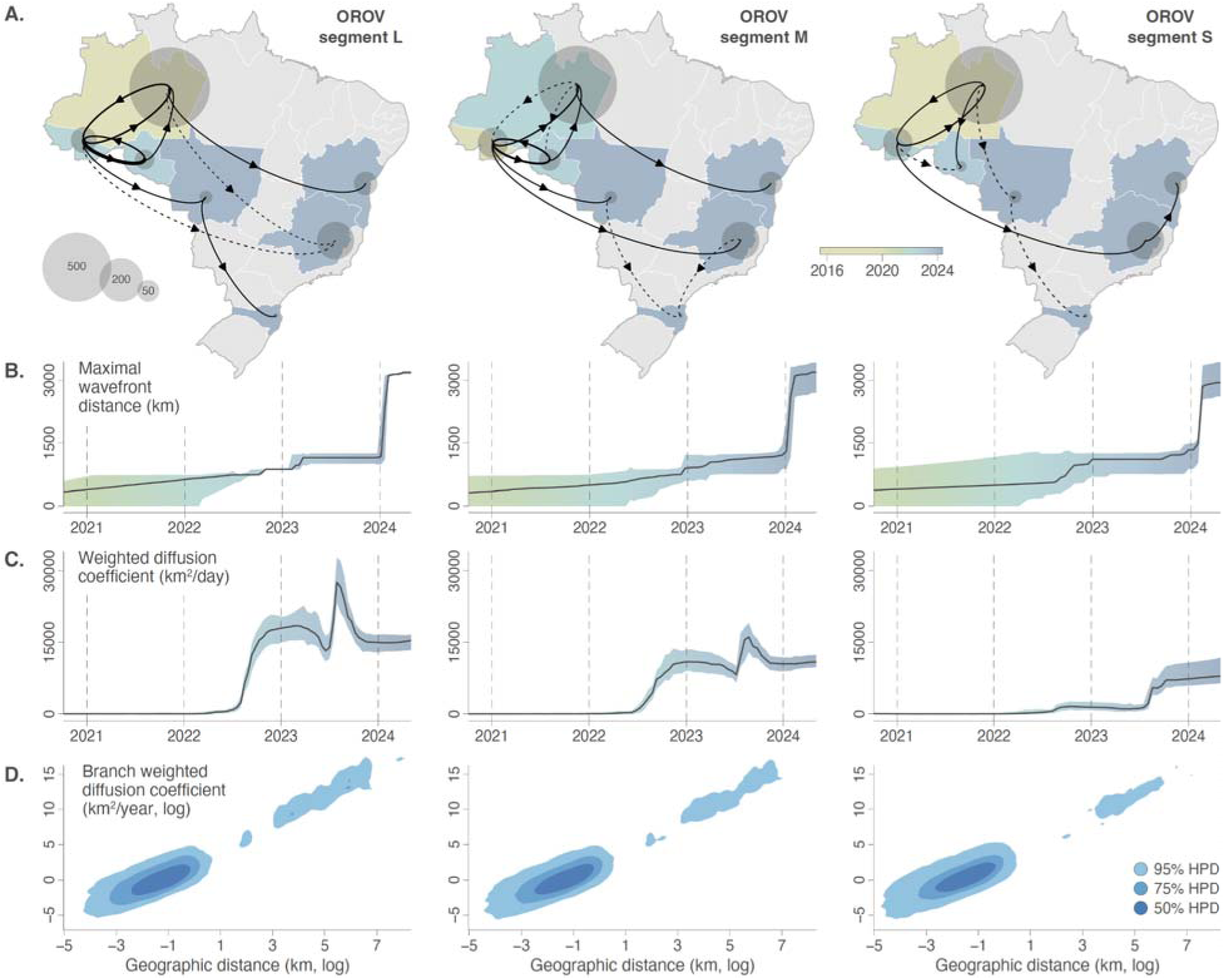
Dispersal history and dynamics of OROV lineages in Brazil. **(A)** Dispersal history of OROV lineages inferred through continuous phylogeographic reconstructions. Lineage dispersal events between Brazilian states with a posterior probability >=0.95 are displayed by solid arrows, and dispersal events with a posterior probability <0.95 are displayed by dashed arrows. Additionally, the location of the different areas is represented by transparent grey dots whose surface is proportional to the number of local lineage dispersal events, i.e. phylogenetic branches inferred as remaining in that state. Brazilian states are coloured according to the estimated date of the first invasion event (median date computed from the 100 trees sampled from the posterior distribution). **(B)** Evolution through time of the spatial wavefront distance, representing the maximal distance from the epidemic origin over time. **(C)** Evolution through time of the weighted diffusion coefficient, a dispersal metric that measures the dispersal capacity of viral lineages. **(D)** Kernel density plots with the branch-weighted diffusion coefficient against the geographic distance travelled by each branch (both axes being log-transformed).

The earliest lineage dispersal events were restricted to the Amazon basin and inferred to be before 2020, with the virus gradually spreading to other Brazilian states in a southeasterly direction in 2023 and 2024. By further examining the spatial dissemination of OROV lineages across Brazil, we found that the virus reached a maximal wavefront distance of over 3,000 km from its epidemic origin through the entire dissemination period (**Figure 1B**). This reconstruction further captured two distinct expansion phases during which the wavefront distance increased rapidly, indicating the invasion of new areas >2024 (**Figure 1B**). The rapid expansion of wavefront distance in 2024 corresponds to the increase in cases in areas outside the Amazon (**Supplementary Figure S1**). Furthermore, diffusion coefficient estimates have evolved through time, with increases in mid-2022 and mid-2023 consistently detected in all three OROV segments (**Figure 1C**). As a result, the weighted diffusion coefficient estimated for the overall period was also notably high: 582 km^2^/day (95% HPD = [477, 672]) for segment L, 574 km^2^/day (95% HPD = [464, 677]) for segment M, and 515 km^2^/day (95% HPD = [435, 675]) for segment S. These values are significantly higher than those reported for key viral dispersal events, such as for West Nile virus spread via birds in North America (*20*), reflecting a substantial dispersal capacity. In 2024, a large number of dispersal events occurred within the states of Mato Grosso, Minas Gerais and Bahia (midwest, southeast and northeast Brazil, respectively), which were previously thought to be non-endemic for OROV transmission.

With respect to dispersal distance and diffusion velocity, we observe a large group of phylogenetic branches associated with relatively short dispersal distance (<20km) and slow diffusion (<4km^2^/day; **Figure 1D**), and a subsequent group of branches that correspond to faster long-distance dispersal events (**Figure 1D**). This supports a multi-scale expansion process with a combination of short-distance diffusive movement and fast long-distance jumps, with some of the latter likely reflecting human-mediated virus movements. We also identified substantial isolation-by-distance (IBD) patterns, with Pearson correlations between patristic and log-transformed geographic distances between samples close or greater than 0.5 for all three segments (0.470 (95% HPD = [0.360, 0.599]) for segment L, 0.575 (95% HPD = [0.286, 0.607]) for segment M, and 0.684 (95% HPD = [0.326, 0.710]) for segment S). Overall, these dispersal metrics provide a clear indication of rapid long-distance dispersal events during the recent OROV expansion in 2024 beyond the Amazon Basin, likely human-mediated, followed by more localised viral circulation. This view is supported by an examination of air travel data from Brazil, with considerable human mobility between the Amazon region and other parts of the country, primarily through the airport in Manaus (**Supplementary Figure S2**). Air travel data also support the inferred long-distance dispersal routes from the state of Acre to the east coast of Brazil, and also highlight the intermediate role played by the state of Mato Grosso in the virus’ migration out of the Amazon region.

### Ecological factors associated with the transmission of OROV in Brazil

To elucidate the ecological factors associated with the spatial expansion of OROV in Brazil, we analysed virus dispersal history in relation to 28 environmental factors (including land-use, climatic, and demographic variables) (**Supplementary Figure S3**). In the absence of established species distribution models for *Culicoides paraensis,* the presumed primary vector for OROV, we also considered the ecological suitability of *Ae. aegypti* as a covariate, as this species could act as a proxy for urban anthropophilic vectors, or as a potential secondary vector for OROV (*21*, *22*). *Aedes* (*Ae.) aegypti* is a well-established vector for several arboviruses, including dengue, Zika, and chikungunya, and its wide distribution across urban areas in Brazil (**Supplementary Figure S3**) may facilitate the spread of OROV, especially in densely populated regions with favourable breeding conditions. While so far *Ae. aegypti* has not been demonstrated as a viable vector, the situation with the novel reassortant lineage could be different (*21*).

Using high-resolution environmental rasters for each covariate (**Supplementary Figure S3**), we extracted environmental values at the geographic location of each phylogenetic node. These correspond to inferred positions through continuous phylogeographic analysis for internal nodes, and to sampling locations (down to municipality level) for tip nodes (**Supplementary Figure S4**). Our analyses revealed that several covariates were temporally associated with OROV dispersal events, highlighting distinct viral circulation environments over time. We assessed shifts in dispersal environments relative to three specific time points: (1) prior to the re-emergence and epidemic expansion in the Amazon (<mid-2023), (2) during the Amazon-restricted transmission phase (<2024), and (3) during the national expansion phase (>2024). When comparing dispersal events before and after 2023-2024, different environmental conditions for OROV circulation become apparent (**Figure 2**). Certain dispersal environments seem to shift at the mid-2023 time point, while others at the 2024 time point (**Figure 2, Supplementary Figure S4**). For instance, prior to 2024, dispersal events occurred on average in areas with relatively higher evergreen broadleaf forest cover, higher precipitation and temperature, but lower population density and cocoa cultivation areas (**Figure 2, Supplementary Figure S4**). Interestingly, most lineage dispersal locations before 2024 were associated with a mean temperature of ∼27°C, which may indicate an optimal viral replication environment for OROV in its vector and is supported by preliminary thermal biology studies on biting midges (*23*). However, dispersal environments were already shifting towards areas with high population density, increased urbanisation, and larger cocoa cultivation areas around mid-2023 (**Figure 2, Supplementary Figure S4**). With strong Bayes factor support (*24*), our analyses demonstrated that these trends in dispersal environments were consistent across the posterior distribution of trees obtained through continuous phylogeographic inference.

**Figure 2.**
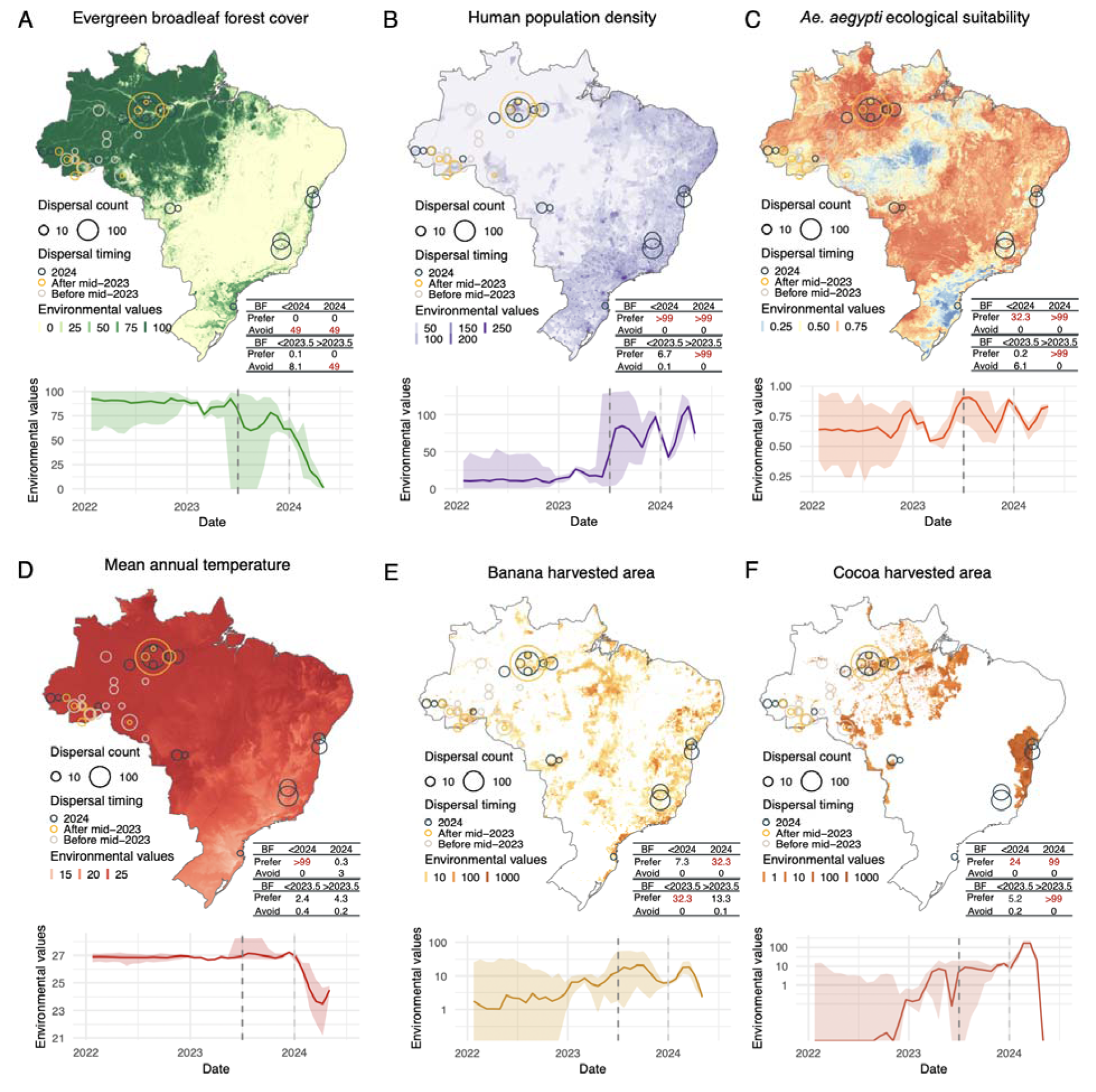
Environmental conditions associated with OROV lineage dispersal locations over time (for segment M). Figure panels show the spatial distribution of six main environmental factors (units specified): evergree broadleaf forest cover (%) **(A)**, human population density (normalised between 0 and 255 per km^2^ for visual clarity) (**B**), *Ae. aegypti* ecological suitability (probability of occurrence) (**C**), mean annual temperature (°C) (**D**), banana harvested area (in hectares, log-transformed) (**E**), and cocoa harvested area (in hectares, log-transformed) (**F**) in the top rows. Circles on the map depict the end node of dispersal locations inferred by continuous phylogeography, sized by the number of dispersal events in an area, and coloured by the timing of the event. Bottom rows of eac figure panel are line graphs depicting the environmental covariates associated with the locations of OROV lineag dispersal events in Brazil. Each plot illustrates how specific ecological conditions have changed over time (2022-2024) at the sites of viral lineage dispersal. The embedded tables show the association between environmental conditions and the dispersal location of inferred OROV lineages. Based on the analysis of 100 posterior trees obtained from continuous phylogeographic inference, the table reports Bayes factor (BF) supports for associatio between environmental raster values and tree node locations. Following the scale of interpretation of Kass and Raftery (*24*), we highlight BF values >20 considered as strong supports.

To statistically test the association of environmental conditions with the spread of OROV lineages, we employed a landscape phylogeographic approach to analyse the environmental values extracted at the tree node positions (*25*). Specifically, for the three distinct time periods mentioned above, we tested whether inferred OROV lineages tended to preferentially circulate in or avoid certain environmental conditions. Statistical support (Bayes factors [BF]) was obtained by comparing the results from phylogeographic reconstructions with a null dispersal model, in which a new continuous diffusion process was randomly simulated along the same tree topologies. During both transmission phases divided by the 2024 cut-off, our results reveal strong support (BF >20 for at least two out of three segments) for preferential circulation of inferred OROV lineages in areas with higher population density and urbanisation, lower evergreen broadleaf forest cover, and areas associated with cocoa cultivation (**Figure 2, Supplementary Table S1**). We also highlight a preferential circulation of inferred OROV lineages in areas associated with banana cultivation in the expansion phase (>2024), as well as in the pre-expansion phase (<2024) for segment L (**Supplementary Figure S6**). These results are consistent with existing knowledge about *C. paraensis* larvae developing in microhabitats of decaying debris from banana and cacao plantations (*26*–*28*). Specific to the Amazon-only transmission phase (<2024), our results also indicate preferred circulation in areas with higher mean annual temperatures (**Figure 2D**). Additionally, our analyses indicate strong support for preferential circulation of inferred OROV lineages in areas associated with higher *Ae. aegypti* ecological suitability, either in both phases (segment M) or in the only expansion phase (segments S and L; **Figure 2B, Supplementary Figure S5, S6**) (**Supplementary Table S1**), although this could be an indirect association with human human density. These results indicate similarities in environmental conditions associated with OROV lineage circulation before and after the 2024 cut-off, suggesting that the expansion of the virus to areas outside the Amazon was not associated with drastically different environmental conditions. However, an examination of the differences in dispersal locations between transmission phases prior to and after mid-2023 reveals compelling differences: for viral lineages inferred prior to mid-2023, there was not strong support for a preferential circulation in densely populated areas, areas associated with a lower evergreen broadleaf forest coverage, and those more ecologically suitable for *Ae. aegypti,* although urbanised areas were still preferred (**Supplementary Table S1)**. This suggests ecological differences underlying endemic OROV circulation within the Amazon before the re-emergence and rapid epidemic expansion at the end of 2023. Overall, our findings reveal similar trends across the three segments analysed (**Supplementary Figure S5, S6, Supplementary Table S1**).

### Mapping ecological niches for OROV transmission and range expansion in Brazil

The increased detection of OROV cases in humans also provides an opportunity to apply modelling to examine areas ecologically suitable for local circulation of the virus, leading to human infections. We used an ensemble modelling approach to reveal OROV transmission suitability across Brazil. The suitability index ranges from 0 (unsuitable conditions) and 1 (highly suitable conditions) and illustrates the potential geographic areas where environmental conditions are most favourable for OROV transmission (**Figure 3A**). The disease presence points used as model input, represent OROV circulation leading to human cases from 450 geocoded sampling locations in Brazil (based on molecular testing and sequencing records) from 1957 to 2024 (∼80% corresponding to 2023-2024). Given this large time span, climatic variables were matched to the corresponding decade of the occurrence point. In the context of presence-only ecological niche modelling, where true disease absence data is unavailable, we sampled pseudo-absence points. Pseudo-absence points sampling was informed by a kernel density estimate of human population density to reflect surveillance efforts which we assumed is proportional to human population density, with an exclusion radius around presence points (**Supplementary Figure S7**). The model incorporated the same environmental covariates used in the landscape phylogeography analyses (**Figure S1**), while testing for overfitting along with model performance (**Supplementary Table S3**). A principal component analysis (PCA) was performed to assess multicollinearity between the environmental covariates (**Supplementary Figure S8**), resulting in a final selection of nine variables: *Ae. aegypti* ecological suitability, annual mean temperature, evergreen broadleaf forest cover, croplands cover, elevation, human population density, annual mean monthly precipitation, as well as cocoa and banana harvested area coverages. We also report variability among our model predictions as a measure of uncertainty (**Supplementary Figure S9**). Uncertainty is higher in the central regions of Brazil (**Supplementary Figure S9**), where lower population density and limited sampling reduce the accuracy of our predictions. Overall, our findings indicate that the highest ecological suitability for local OROV circulation (index >0.7) is concentrated in the northern regions (**Figure 3A**), particularly within the Amazon Basin, which aligns with previous studies identifying this area as a significant epicentre for the virus (*10, 11*). Additionally, moderate to high suitability areas extend toward the northeast and central-western parts of Brazil, particularly in states such as Para, Maranhao, Bahia, and Mato Grosso, suggesting an expansion into previously non-endemic regions. Our model also highlights the potential for OROV transmission to move beyond traditional sylvatic cycles, affecting urban and peri-urban areas, particularly in the northeast (**Figure 3A**), where human interaction with vectors may amplify transmission risks. This trend is especially notable for highly suitable areas around the coast of Brazil, where over half of the 200 million people in Brazil reside (**Figure 2B**).

**Figure 3.**
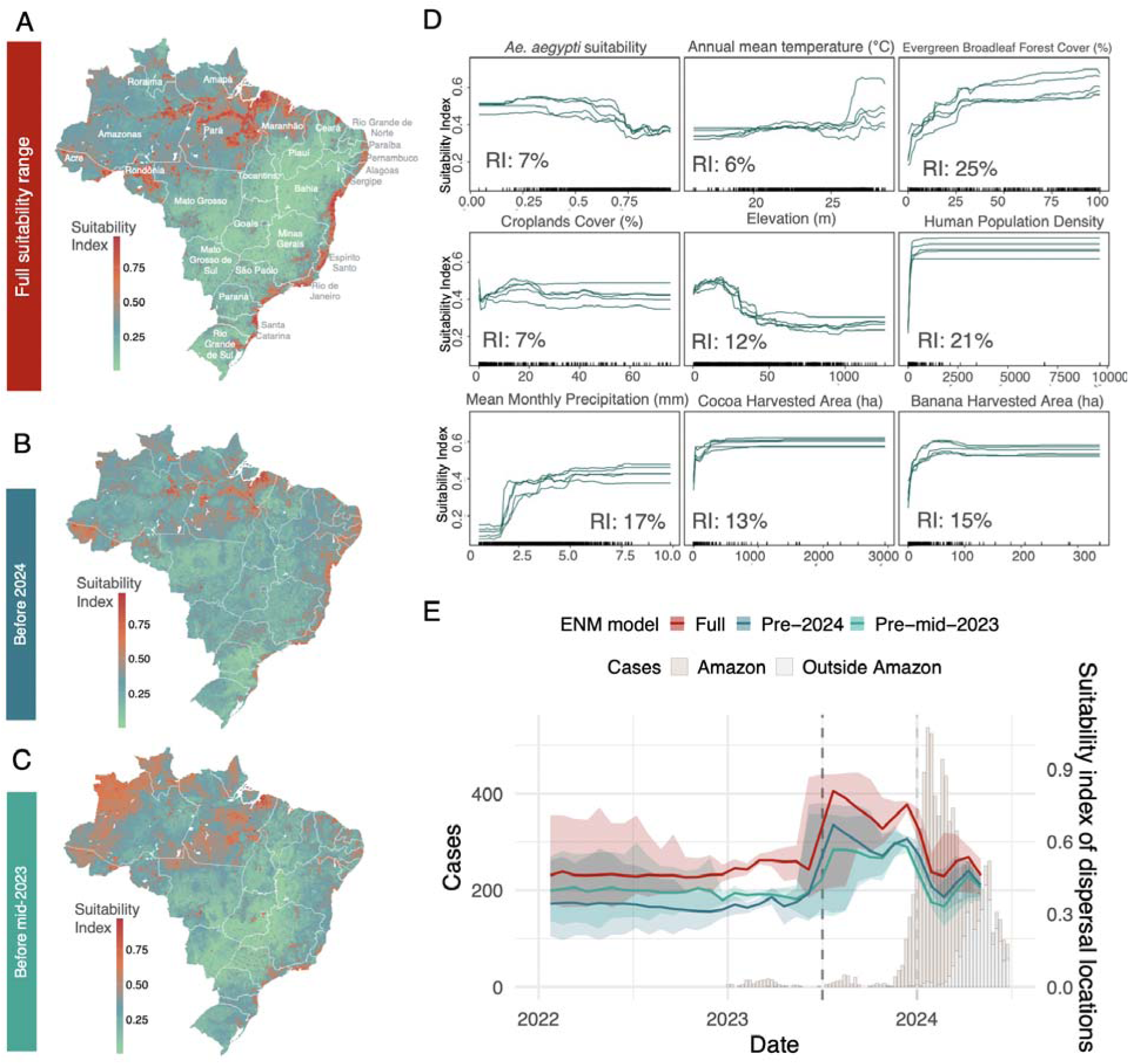
Ecological niche prediction for OROV local circulation in Brazil. A) Predicted ecological suitability for OROV transmission across Brazil utilising all disease occurrence points. Suitability predictions range from unsuitable (0) to highly suitable (1). B) Ecological suitability prediction using input disease occurrence points sampled before 2024. C) Ecological suitability prediction using input disease occurrence points sampled before mid-2023. D) Response curves and relative importance (RI) for individual environmental factors obtained from the random forest (RF) suitability prediction model. These response curves (five iterations) depict the relationshi between the environmental factors and the response (the ecological suitability of OROV transmission). E) Ecological suitability values of OROV dispersal locations (for segment M) overlayed on weekly recorded OROV cases in Brazil. Suitability values are estimated from three ecological niche models (ENM), as described in the text.

To determine the individual contributions of each environmental factor to our suitability prediction, we further calculated their relative importance (RI) in the resulting ecological niche models. We found that evergreen broadleaf forest cover and human population density contributed the most to the model predictions, followed by precipitation, and banana and cocoa agricultural lands (**Figure 3D**). This aligns with factors identified in the independent landscape phylogeographic analyses, and provides important insights into the disease ecology of OROV. We plotted response curves to assess the relationship between the environmental factors and OROV transmission suitability. These curves illustrate how ecological suitability varies with changes in one factor while all others are kept constant at their mean. We observe a clear tipping point in the suitability index with mean monthly precipitation, where environments with <2.5 mm mean monthly precipitation appear unsuitable for OROV transmission (ecological suitability ∼0) whereas suitability increases considerably above this threshold (ecological suitability >0.4) (**Figure 3D**). For temperature, there is a clear increase in OROV suitability in environments with annual mean temperatures above 25°C (**Figure 3D**). OROV transmission suitability decreases considerably at high elevations, while higher human population density, evergreen broadleaf forest cover, and higher areas of banana and cocoa agriculture appear to favour transmission (**Figure 3D**).

To investigate whether the expansion of OROV in Brazil in late 2023 and 2024 was associated with an expansion of its ecological niche, we compared ecological niche models trained on pre-mid-2023, pre-2024, and all available occurrence records. Specifically, we compared both the resulting maps of OROV ecological suitability and the capacity of each category of models to predict the distribution of most recent occurrence data. The suitability range obtained with the pre-2024 models (**Figure 3B**) highlights a similar spatial distribution of highly suitable regions compared to that obtained with the full models (**Figure 3A**). While the full suitability estimates show a slight expansion of suitable areas, the pre-2024 model was able to predict the 2024 occurrence points with a relatively high performance (TSS = 0.6, AUC = 0.862; **Supplementary Table S2**). However, the pre-mid-2023 suitability range (**Figure 3C**) shows clear differences from the full model, particularly in the landscape of ecological suitability within the Amazon; and the models had a lower predictive ability for occurrence points sampled after mid-2023 (TSS = 0.4, AUC = 0.55; **Supplementary Table S2**). This indicates a shift in ecological suitability after mid-2023, associated with OROV amplification within the Amazon, prior to circulation in other parts of the country. Given that this shift did not occur in 2024, viral circulation outside the Amazon was likely associated with OROV lineages recently reaching areas that were already ecologically suitable for local OROV transmission. This is supported by our earlier findings of long-distance and rapid dispersal events from continuous phylogeography.

Integrating results from the ecological niche modelling with the inferred dispersal histories, we can further assess the estimated ecological suitability values at each dispersal location associated with OROV circulation across space and time (**Figure 3E**). Examining dispersal locations occurring prior to mid-2023, after mid-2023, and in 2024, it becomes evident that for most of its dispersal history, OROV circulated in regions of moderate ecological suitability (∼0.4-0.5), consistent across the three temporal models we estimated. This was then followed by a peak in ecological suitability values, reaching ∼0.6-0.8 associated with OROV dispersal from mid to late 2023, coinciding with unprecedented epidemic expansion within the Amazon (**Figure 3E, Supplementary Figure S1**). This peak in ecological suitability was likely due to the virus being introduced to areas of the Amazon that had a combination of favourable environments and higher population densities, such as Manaus, leading to an amplification of transmission. This expansion in a highly suitable and densely populated environment, in turn, likely facilitated the pathogen’s spread beyond its usual transmission range. Additionally, this amplification environment was more connected via human mobility to the rest of the country (**Supplementary Figure S2**), compared to more remote areas of the Amazon, where the virus had previously circulated. This further emphasises the need for improved surveillance in blind spot regions where transmission suitability is high, but where cases may be underreported or genomic data may be scarce. In such regions, introductions could rapidly lead to amplified outbreaks. A subsequent drop in ecological suitability associated with dispersal locations after 2024 is likely due to the virus colonising areas with less favourable environmental conditions (e.g., lower mean annual temperatures - **Figure 2D**, higher elevation, and lower precipitation - **Supplementary Figure S4**), despite high population densities.

### Discussion, limitations and conclusions

The recent emergence and expansion of OROV into previously non-endemic regions underscores the critical need for a deeper understanding of the factors associated with its spread. In response to the 2023-2024 outbreaks, recent studies have described the emergence of a reassortant lineage (*10*, *11*), investigated spatiotemporal movement dynamics at a smaller scale (*6*, *7*, *10*, *11*), reported severe clinical outcomes (*9*), and documented biological differences in the novel lineage (*29*). However, none had investigated the ecological mechanisms associated with the amplification of OROV within the Amazon and in other parts of the country (*30*). Here, we integrated phylogeographic and ecological modelling approaches to explore potential correlates of OROV transmission and range expansion. Our phylogeographic reconstructions provide initial evidence for the underlying dynamics governing OROV’s amplification and spread in Brazil. By analysing environmental phylogeographic relationships and the ecological niche associated with viral transmission during distinct phases, we found that OROV circulated in areas of enhanced ecological suitability immediately preceding its explosive expansion in the Amazon. Our phylogeographic reconstructions also demonstrate that this expansion occurred with a considerable diffusion capacity, characterised by a series of rapid long-distance dispersal, most likely human-mediated through air travel across the country.

Environmental conditions play a pivotal role in shaping OROV transmission dynamics (*31*). Zones undergoing land-cover transitions, particularly those involving deforestation and agricultural activities, have emerged as critical hotspots for virus spread (*32*, *33*). As these areas transition from sylvatic (forest) environments to more urbanised or agricultural landscapes, the resulting habitat changes bring vectors and reservoir hosts into closer contact with human populations, creating new opportunities for OROV transmission, and these must be studied using an integrated approach (*34*). Through the landscape phylogeographic and ecological niche modelling analyses conducted in this study, we propose a mechanism for the expansion of OROV, first within and then beyond the Amazon. We detect both a shift in estimated ecological suitability and differences in the environmental space visited by dispersal events prior to epidemic amplification within the Amazon. However, this was not concurrent with the emerging circulation in regions outside the Amazon, where the virus has not widely circulated before. This consistently supports a two-step expansion process, in which circulation within a highly suitable environment led to an explosive outbreak in the Amazon, which then facilitated OROV’s spread beyond its usual transmission range. These findings are supported by independent reconstructions of all three genomic segments of the virus. While previous studies have identified the Amazon region as the primary source of OROV emergence in Brazil (*10*, *11*), the re-emergence or introduction of the virus in the vicinity of Manaus, the capital of the vast state of the Amazon, would have exposed a large number of naive human hosts and contributed to an amplified outbreak. Higher suitability in the amplification zone within the Amazon could therefore be attributed to a combination of favourable environmental conditions and densely populated urban areas. Our results also suggest that the novel circulation of OROV in non-endemic parts of Brazil, albeit concerning, could be self-limiting, particularly in areas with less favourable environments.

Our study illustrates the power of integrating phylodynamics, landscape phylogeography and ecological niche modelling in elucidating complex eco-epidemiological dynamics of re-emerging arboviruses and urban amplification. These findings have several implications. First, by using complementary analytical methods and a comprehensive genomic data set, we identify key environmental factors important for virus transmission. Results from both approaches implicate human population density, banana and cocoa cultivation, and temperature as factors associated with favourable transmission environments. Such information can directly inform public health planning and mitigation measures, such as vector control activities around banana and cocoa plantations, particularly near urban areas. A recent study also implicated banana and cocoa as important crop types in epidemic locations (*35*). Second, while not a confirmation, our work provides a robust initial indication that OROV expanded opportunistically by finding more favourable environments for transmission, which were also more connected to the rest of the country. Other hypotheses have explored the idea that higher viral replication in mammalian cells could allow more efficient transmission to vector and onwards, or that the immune escape ability of the new reassortant lineage may play a role (*29*). Synthesising recent work with our findings identifies a proposed mechanism for this emerging outbreak. It is possible that genetic reassortment resulted in a viral genotype characterised by more efficient infection and transmission between humans and the vector, resulting in higher viremia, in turn explaining the epidemic expansion in favourable urban environments detected in our study. The approach we propose is highly adaptable to risk mapping and the characterisation of disease ecology for other arboviruses and zoonotic infections. Finally, the concentration of high-suitability areas in Brazil’s coastal regions is concerning, as this is where most of the population resides and where multiple arboviruses already circulate. Multiple introduction events into these areas could lead to further amplification of this pathogen, particularly given the presence of immunologically naive populations, or the circulation of a variant that can evade prior immunity (*29*). Additionally, several suitable areas, especially in the northeastern and central-west regions, remain surveillance blindspots for OROV. These areas may harbour undetected transmission due to limited sampling. Prioritising these blind spots for active surveillance is essential for timely viral detection and early interventions to prevent outbreaks.

This study needs to be interpreted in light of certain limitations. Our landscape phylogeographic approach is affected by the pattern of the sampling effort, as approximately half of the node locations are sequenced tips, making them prone to sampling biases (*25*). This means that this approach cannot currently test the true drivers of transmission but rather test the strength of associations of dispersal environments. Additionally, we lack a species distribution model for the actual vector, *Culicoides paraensis*, and vertebrate host species that typically compose OROV’s natural reservoir, which limits our ability to fully understand host-vector-specific environmental suitability. To mitigate this bias, we test associations against a null dispersal model generated through simulations, and only consider factors with the strongest Bayes factor support (>20). Additionally, this study was designed only to examine the ecological factors associated with epidemic expansion in Brazil. The potentially critical role of reassortants in viral adaptation and fitness has not been addressed here. Our phylogeographic analyses focused solely on the reassortant lineage that emerged in Brazil. To elucidate the role of these evolutionary processes in the virus’ ability to adapt to new niches, hosts or vectors, our approach could be extended to analyse evolutionarily distinct lineages of OROV that previously circulated in Brazil or neighbouring regions. The role of vectors is central to the expansion of OROV, with the primary vector *Culicoides paraensis* likely adapting to new landscapes. The interaction between the virus, its vectors and its reservoir hosts is crucial for both the colonisation of new transmission zones and the maintenance of transmission in established areas. Our analysis also revealed increased exposure of OROV to urban and peri-urban vectors such as *Ae. aegypti,* which could facilitate its spread in densely populated regions. However, our approach cannot definitively support that this vector was directly involved in epidemic expansion. While the virus has previously been detected in *Ae. aegypti* mosquitoes (*21*), further vector competence studies, particularly against this novel reassortant lineage, are required to understand the risks posed by increased exposure of this virus to secondary urban vectors.

The geographic expansion of OROV into new regions highlights the pressing need for integrating environmental monitoring into public health frameworks. To effectively predict and mitigate the risks posed by OROV and other arboviruses, surveillance efforts must account for the complex interaction between environmental changes, vector ecology and human behaviour. As OROV continues to adapt to new ecological niches, driven by combinations of genetic evolution and both natural and anthropogenic factors, a deep understanding of these dynamics will be essential for developing targeted intervention strategies to control its spread and minimise its public health impact.

## Data Availability

All genomic and disease occurrence data used in this study were already openly available before this study, and analysis scripts will be made openly available via an indexed repository by the time of final publication. During the peer-review process, this is made available on a private repository.

## Acknowledgements

The authors would like to acknowledge Felipe G. Naveca for the diagnostic-based approach developed, which enabled the detection of OROV across Brazil and the Americas.

## Author contribution

Conceptualization: HT., TdO.; Methodology: HT., SD., CM., GD., VF., MST., MD., LCVF., CdE., MM., EW., ECH., CB., RL., MUGK., JL., LCJA., TdO., MG.; Investigation: HT., SD., JP., CM., VF., JL., MG.; Data curation: HT., SD., JM., MG.; Original draft preparation: HT., SD., JP., CM., JL., MUGK., TdO., MG; Visualisation: HT., SD., JP.; Funding acquisition: TdO, LCJA, MG. All authors have read and agreed to the published version of the manuscript.

## Materials and Methods

### Oropouche Virus genomic data

Complete genome sequences of the S, M, and L segments of the Oropouche virus (OROV), obtained from the first extra-Amazon OROV cases reported in the states of Bahia (Northeast Brazil), Minas Gerais (Southeast Brazil), Mato Grosso (Midwest Brazil), and Paraná (South Brazil), were combined with the corresponding segments of recently published full-length OROV sequences belonging to the Brazilian 2022-2024 sublineage (*10*). The OROV sequences used in this study correspond to the Genbank accession IDs: PQ168520-PQ247806 and PP153945-PQ065491. Sequence alignment for each segment (n=545) was performed using MAFFT (*36*, *37*) and subsequently curated manually to remove artefacts using AliView (*38*). Genomic regions identified by RDP5 to have likely been acquired by recombination and genomic segments identified by RDP5 to have been acquired by reassortment, were stripped from the full genome data sets by replacing these regions with gap characters (“-”) in the alignment file, thereby a yielding a free full genome alignment free of recombination and reassortment as previously described (*10*). Sequences with recombination and reassortment signals along the majority of the genome were completely discarded (n=43 for segment S and n=1 for segment L).

### Disease occurrence data

Disease occurrence data was compiled from multiple sources. Epidemiological data on Oropouche virus (OROV) cases were retrieved from the Brazilian Ministry of Health, accessible at https://www.gov.br/saude/pt-br/assuntos/saude-de-a-a-z/o/oropouche. The data set includes confirmed case reports from all Brazilian states where OROV cases have been notified, including but not limited to Acre (AC), Alagoas (AL), Amazonas (AM), Bahia (BA), Ceará (CE), Minas Gerais (MG), Pará (PA), Rio de Janeiro (RJ), and São Paulo (SP). The data cover the years 2023 and 2024, organised by epidemiological week, and include key variables such as the municipality, state, year of occurrence, and the corresponding epidemiological week of reporting. This data was geocoded at the municipality level and occurrence deduplicated by month. Additional OROV occurrence data was gathered and geocoded from all records on the GBIF (years: 1957-present) and Genbank databases (years: 2015-present). After deduplicating all occurrence records, we obtained a total of 450 unique sampling location points covering the years 1957 to 2024, with a majority (∼85%) sampled in 2023-2024.

### Geospatial data

We tested several environmental factors both as associations with viral dispersal locations and as covariates in our ecological niche model. These factors included human population density, main land cover and climatic variables within the study area (Brazil). Each environmental factor was described by a raster that defines its spatial heterogeneity. **Supplementary Table S3** details the source and resolution of each original raster file. Each raster was cropped to match our study area (Brazil) by using Brazil shapefiles from the “rnaturalearth” package in R.

### Air travel data

We retrieved domestic flight data from Brazil’s Civil Aviation Agency (ANAC) using the “flightsbr” R package (Version 0.1.0). This dataset contains information on flight routes, departure and arrival times, airlines, and airport locations. With flight data only available through 2020, we chose to use 2019 as it represents the most recent complete year before the COVID-19 pandemic. It provides a reliable baseline, with reports confirming the recovery of air travel to pre-pandemic levels in 2023 (*39*). We merged the flight data with airport location information, filtering specifically for public airports across Brazil. We focused on creating a network of departing flights, visualised spatially by airport, to highlight regional connectivity patterns. All data processing and visualisations were conducted using R, with geospatial mapping performed using the ggplot2 and sp packages.

### Phylogeographic reconstruction and dispersal statistics

To model the spatiotemporal spread of OROV using spatially-explicit phylogeographic reconstruction using the continuous diffusion model implemented in the software package BEAST 1.10 (*40*), we utilised three different data subsets containing the 2022-2024 OROV sequences from various regions of Brazil of the S (*n* = 501), M (*n* = 545), and L (*n* = 544) segments. Before conducting the phylogeographic analyses, we assessed the strength of the molecular clock signal in each data subset using the root-to-tip regression method available in TempEst v1.5.3 (*41*). Preliminary BEAST reconstructions revealed an outgroup of sequences which diverged from the main clade ∼60 years ago for each segment; these were subsequently discarded from our analyses. Temporal structure was accepted for all datasets as the correlation coefficients were all close to or above 0.5 (S: 0.4972, M: 0.635, L: 0.5123). We reconstructed the spread of OROV lineages within Brazil by using a flexible relaxed random walk diffusion model (*42*), which accommodates branch-specific variation in dispersal rates, with a Cauchy distribution and a jitter window size of 0.01 (*43*). The latitude and longitude coordinates of each sample were used in this analysis. MCMC analyses were run in BEAST v1.10.4, with chains of up to 1 billion iterations each, sampling every 100,000 steps in the chain. The chains were stopped when convergence was reached following the removal of burn-in states. Convergence of each run was assessed using Tracer v1.7.1, ensuring that the effective sample size (ESS) for all relevant model parameters was >200 (*44*). Maximum clade credibility trees were summarised using TreeAnnotator after discarding burn-in samples, the number of which was also determined in Tracer. Finally, the R package “seraphim” (*45*) was employed to extract and map the spatiotemporal information embedded in the posterior trees. We further used “seraphim” to estimate three dispersal statistics from these movement vectors for each segment: maximal wavefront distances, weighted diffusion coefficients (*46*), measuring the dispersal capacity of viral lineages, and an isolation-by-distance (IBD) signal measured as the Pearson correlation between the patristic and log-transformed geographic distances computed for each pair of tip nodes (*20*).

### Landscape phylogeographic analyses

To test the association between environmental conditions (**Supplementary Figure S2**) and dispersal locations of inferred OROV lineages, we employed a landscape phylogeographic approach (*25*). We first extracted and visualised the environmental values explored by phylogenetic branches using the “spreadValues” function implemented in the R package “seraphim”. For these analyses, we sampled 100 posterior trees obtained from the continuous phylogeographic inference. For each posterior tree sampled during the phylogeographic analysis, this function extracts then averages the environmental values at the tree node positions. For each analysed environmental factor, we then obtained a posterior distribution of mean environmental values at tree node positions for each segment data set. To assess the tendency of inferred viral lineages to preferentially circulate within or avoid circulating in specific environmental conditions, we compared the distribution of mean environmental values extracted at node positions in inferred trees (*E*_estimated_) with those extracted at node positions in trees whose dispersal history had been re-simulated under a null dispersal model (*E*_simulated_). To generate such a null dispersal model, a RRW diffusion process was simulated along each tree topology used for the phylogeographic analyses. These RRW simulations were performed using the “simulatorRRW1” function of the R package “seraphim” from the sampled precision matrix parameters estimated by the phylogeographic analyses. Therefore, from these simulations, values at node positions (*E*_simulated_) constitute the distribution of mean environmental values explored under a dispersal scenario that is not impacted by any underlying environmental condition. For each environmental factor and segment-specific phylogeographic reconstruction, we then compared the distribution of *E*_estimated_ values computed from posterior trees with the distribution of *E*_simulated_ values retrieved from the same tree topologies along which a RRW diffusion process had been re-simulated. Specifically, we approximated a Bayes factor (BF) support equal to (*p_e_*/(1- *p_e_*))/(0.5/(1-0.5)). To test if viral lineages tended to avoid circulating within a particular environmental factor *e*, *p_e_* was defined as the frequency at which *E*_estimated_ < *E*_simulated_; and to test if viral lineages tended to preferentially circulate within a particular environmental factor *e*, *p_e_* was defined as the frequency at which *E*_simulated_ < *E*_estimated_. Following the scale of interpretation of Kass and Raftery (*24*), we here highlight BF values >20 considered as strong supports.

### Ecological Niche Modelling

Ecological niche models (ENMs) are built using a variety of statistical methods, each varying in complexity and underlying assumptions about the interaction between species occurrences and environmental factors (*47*). Recent studies have shown that disparities among different model structures can be very large, making model selection difficult (*48*). An alternative is to use an ensemble of models to avoid selecting one single best model but instead to use a group of methods for inference. In other words, the presence of a species might be well classified by some models and misclassified by others, such that making use of an ensemble model can reduce the predictive uncertainty of a single model by combining predictions (*49*). In this study, we applied this approach to model the distribution of OROV transmission by creating a suitability map based on the occurrence of OROV disease in Brazil and relevant environmental variables (see Geospatial Data section). For this analysis, we aggregate all raster maps to the lowest resolution available (∼27km^2^). We used an ensemble of seven statistical, machine learning, and envelope models: Generalised Linear Model (GLM), Generalised Additive Model (GAM), Boosted Regression Trees (BRT), Random Forest (RF), Classification Tree Analysis (CTA), Surface Range Envelope (SRE) and Maximum Entropy Model (MAXENT). To assess the potential expansion of OROV’s ecological niche in Brazil during late 2023 and 2024, we computed and compared ecological niche models (ENMs) using occurrence data from different time periods. Specifically, we created three ensemble models using pre-mid-2023 (89 occurrences), pre-2024 (133 occurrences), and all available occurrence records (450 occurrences).

To assess the models’ performance we use block cross validation (*50*). This is a spatially explicit method used to assess model performance by dividing the study area into geographic blocks. Instead of randomly splitting data, this technique ensures that training and testing data are spatially independent, reducing the risk of spatial autocorrelation. True Skills Statistics (TSS) and receiver-operating characteristic (ROC) curve (area under curve - AUC) are then used to evaluate the predictive performance of the models based on the test (validation) dataset. TSS is equivalent to sensitivity+specificity-1 and ranges from -1 to 1; value of 1 indicates perfect classification, 0 means the model is no better than random guessing, and negative TSS indicates the model performs worse than random guessing. AUC ranges from 0 to 1; AUC of 1 indicates perfect model performance, 0.5 indicates no discrimination (i.e., the model is no better than random guessing), and <0.5 indicates the model performs worse than random guessing. We only retained models with a TSS score of >0.7 to build the ensemble model (**Supplementary Figure S10**). The mean probabilities from each model were then computed, and we weighted the predictions of each model according to its performance during training, giving more weight to better-performing models. The weights ensure that higher-quality models contribute more to the final ensemble prediction. The different resulting ‘suitability indexes’ are then combined to get a single value per site. Once the ensemble predictions are generated, the ensemble model itself is evaluated using the same metrics applied to the individual models; TSS and AUC.

Disease presence points used as input represent OROV circulation occurrence from 450 unique sampling locations in Brazil (molecular testing and sequencing records) from the years 1957 to 2024. Occurrence points with available collection dates were matched to corresponding climatic variables by month. Specifically, temperature and precipitation data for each point were extracted from the monthly climate layers matching the collection month. The sampling of pseudo-absences was done at a 1:1 ratio with presence points and based on those distribution of presence points and a human population density kernel density estimate. Our aim was to sample absences in proportion to the rate of presence points while giving higher priority to areas with greater population density, ensuring more focused sampling in regions where human populations are denser. This approach helps us account for disease testing biases in more urbanized areas in the pseudo-absence distribution. Pseudo-absences were also selected within a perimeter of 50-300 km around presence points, ensuring that absence points are neither too close to presence points (to avoid the same niche) or too far (to promote localised sampling strategy).

To determine the independent contributions of each variable to our suitability prediction, we further calculated their relative importance (RI). In the case of random forest (RF) models, RI is computed by assessing how frequently a variable is selected for splitting at tree nodes, weighted by the squared improvement in model performance resulting from each split, and averaged across all trees (*51*). Higher RI values indicate greater relative contribution of that variable to the predictive performance of the model. We also produced response curves to visualise the effect of each variable on suitability predictions in the RF models. These response curves allow us to observe how changes in a single variable influence the predicted outcome, while other variables are held constant (at their mean). By examining these relationships, we gain insights into how each variable individually contributes to the model’s overall predictions.

## Supplementary Figures

**Figure S1.**
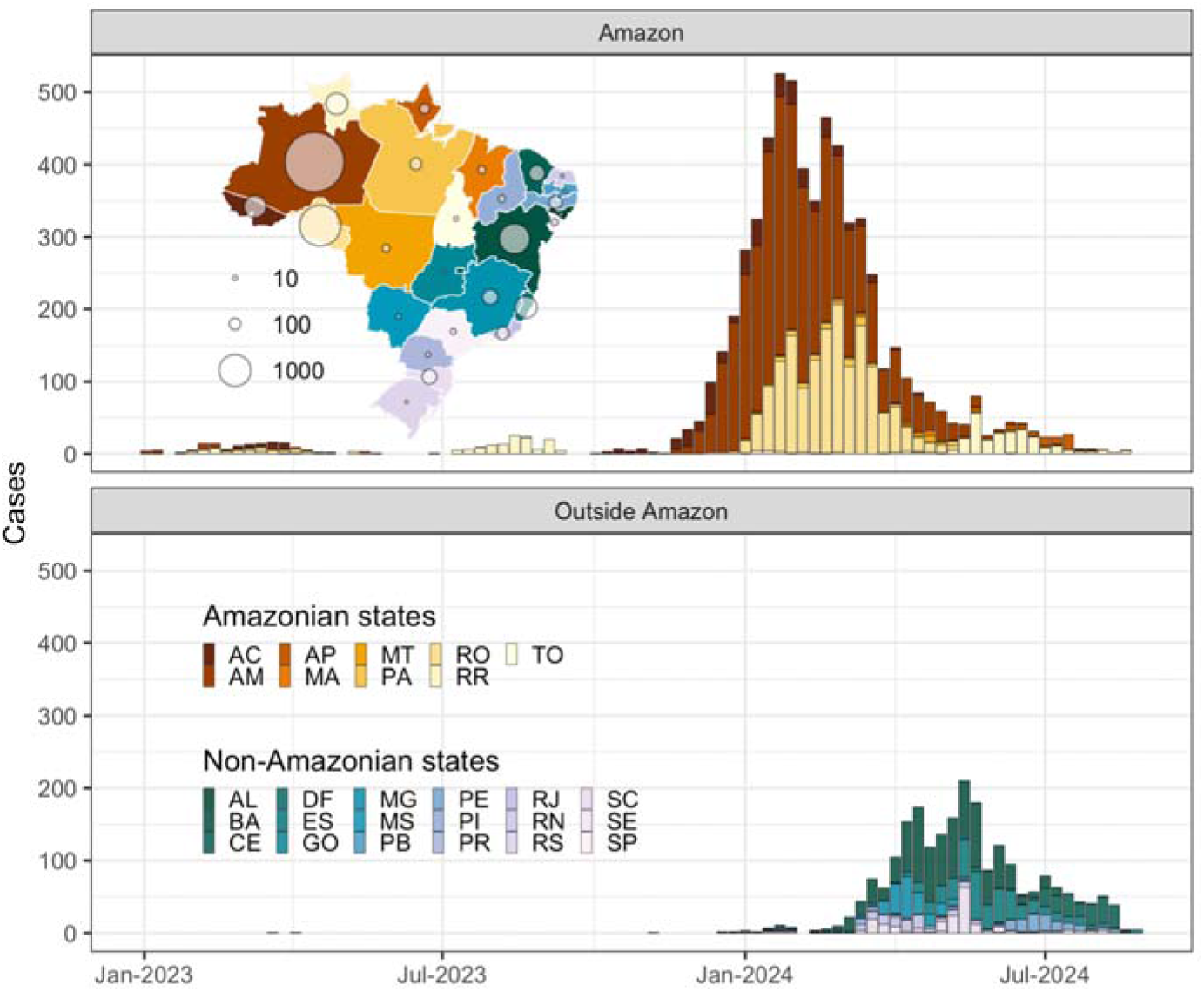
Epidemiological curve of OROV cases in Brazil. Weekly cases are shown for 2023 and 2024 divided into two epidemiological curves, one for states in the Amazon region, and one for states outside the Amazon region. The inset map is coloured by the specific state, and the circles represent the total number of recorded OROV cases i that state.

**Figure S2.**
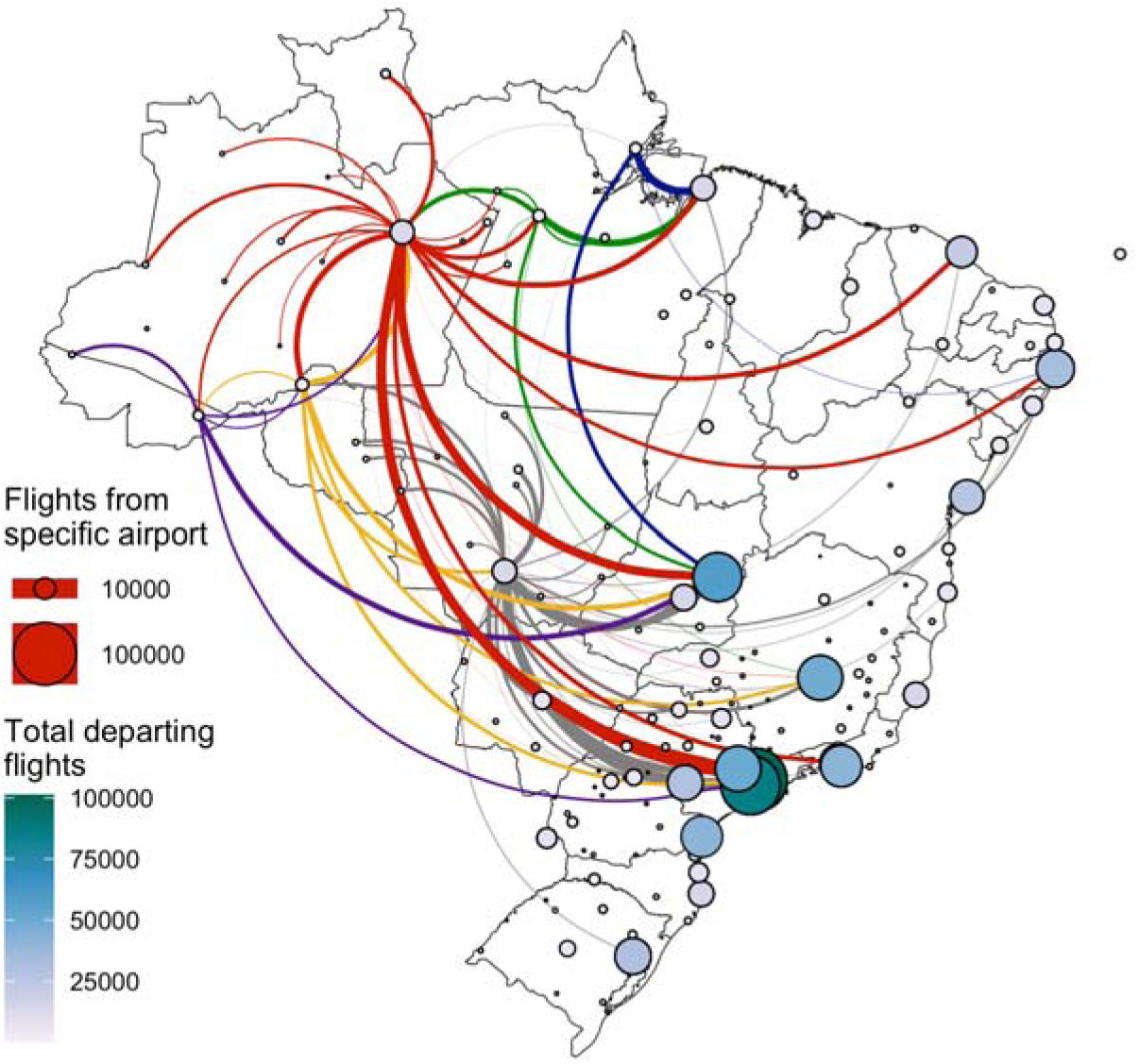
Human mobility through air travel in Brazil. The figure captures air travel data in Brazil in 2019. The map shows the total number of departing flights from all airports in Brazil. Circles are both coloured and sized by the number of flights departing from an airport origin location. The coloured curves show the number and network of flights from airports of specific municipalities, namely Manaus in state of Amazonas (red), Santarém in the state of Pará (green), Cuiabá in the state of Mato Grosso (grey), Porto Velho in the state of Rondônia (yellow), Rio Branco in the state of Acre (purple), and Macapá in the state of Amapá (blue).

**Figure S3.**
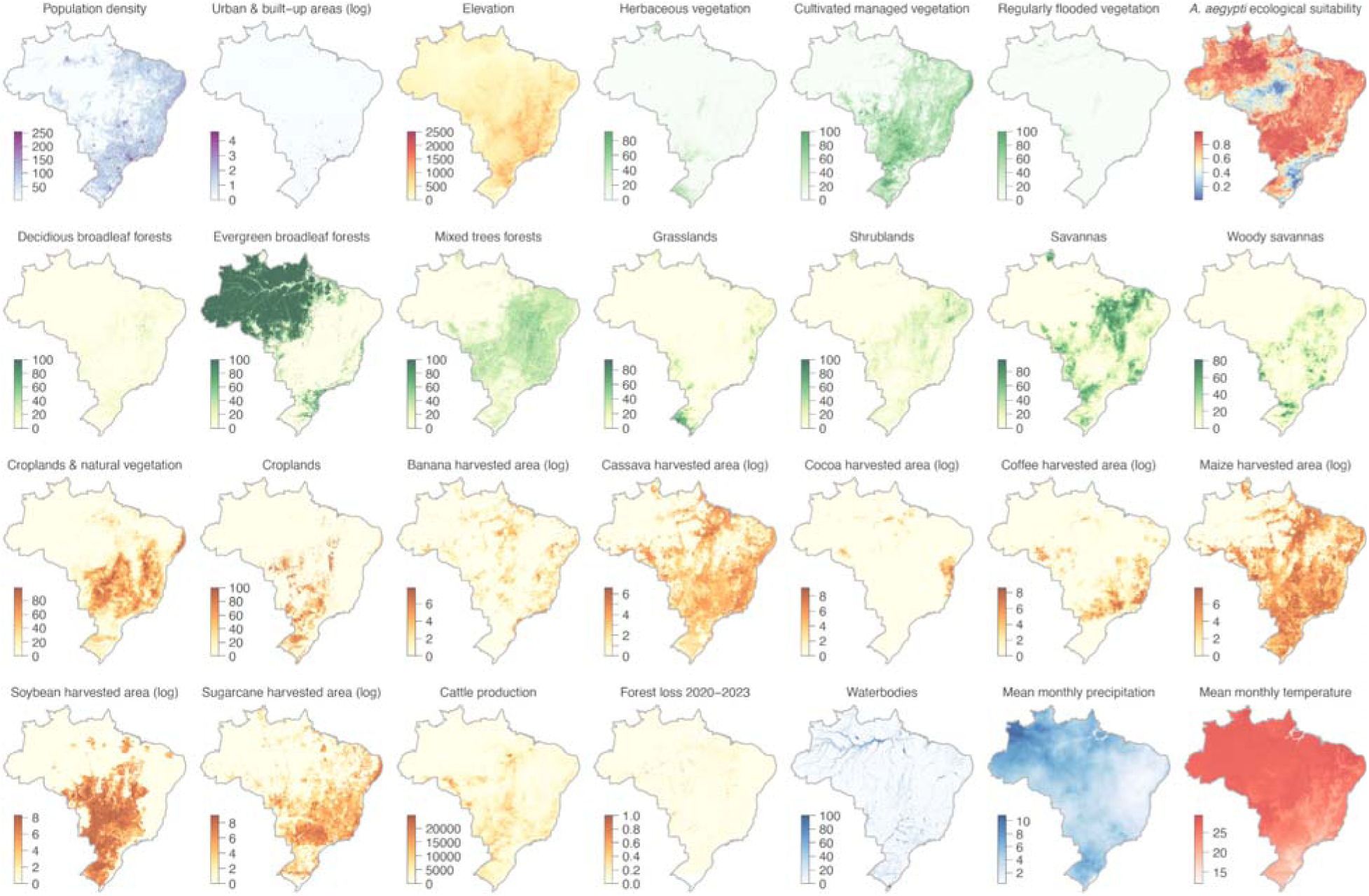
Environmental covariates analyzed in the context of the range expansion of OROV in Brazil. Various environmental rasters, such as demographic, land-use, and climatic covariates, were analyzed in the study t investigate their association with the spread of the OROV in Brazil. Demographic variables encompass populatio density and urban areas, while land-use patterns focus on the presence of croplands, water bodies, and regions impacted by deforestation linked to specific agricultural activities, such as cocoa, soy, and banana crops.

**Figure S4.**
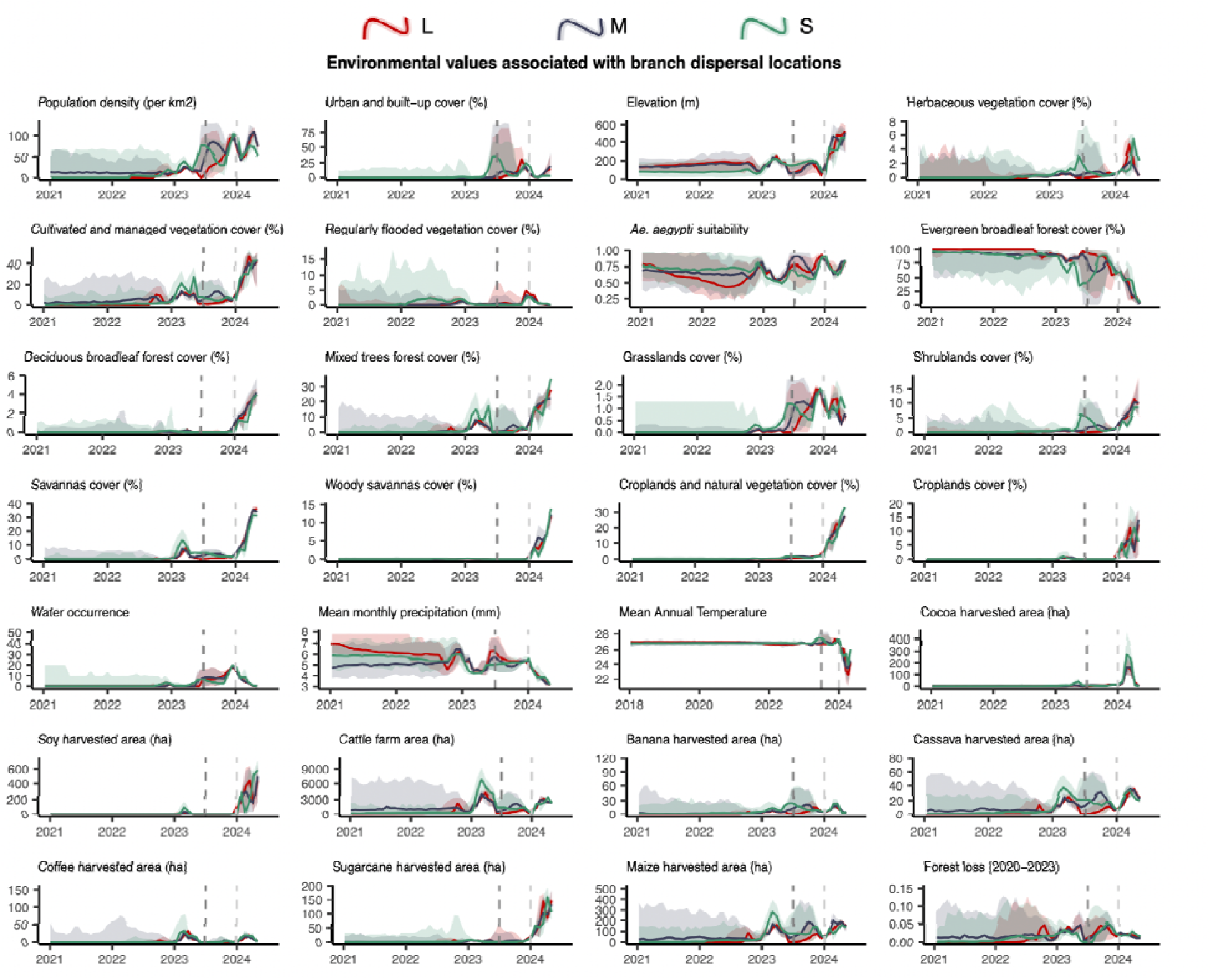
Environmental values associated with OROV branch dispersal locations over time. Line graphs depicting the environmental covariates associated with the locations of OROV lineage dispersal events in Brazil. Each plot illustrates how specific ecological conditions have changed over time (2021-2024) at the sites of viral lineage dispersal. This is shown for segment L (in red), segment M (in blue), and segment S (in green).

**Figure S5.**
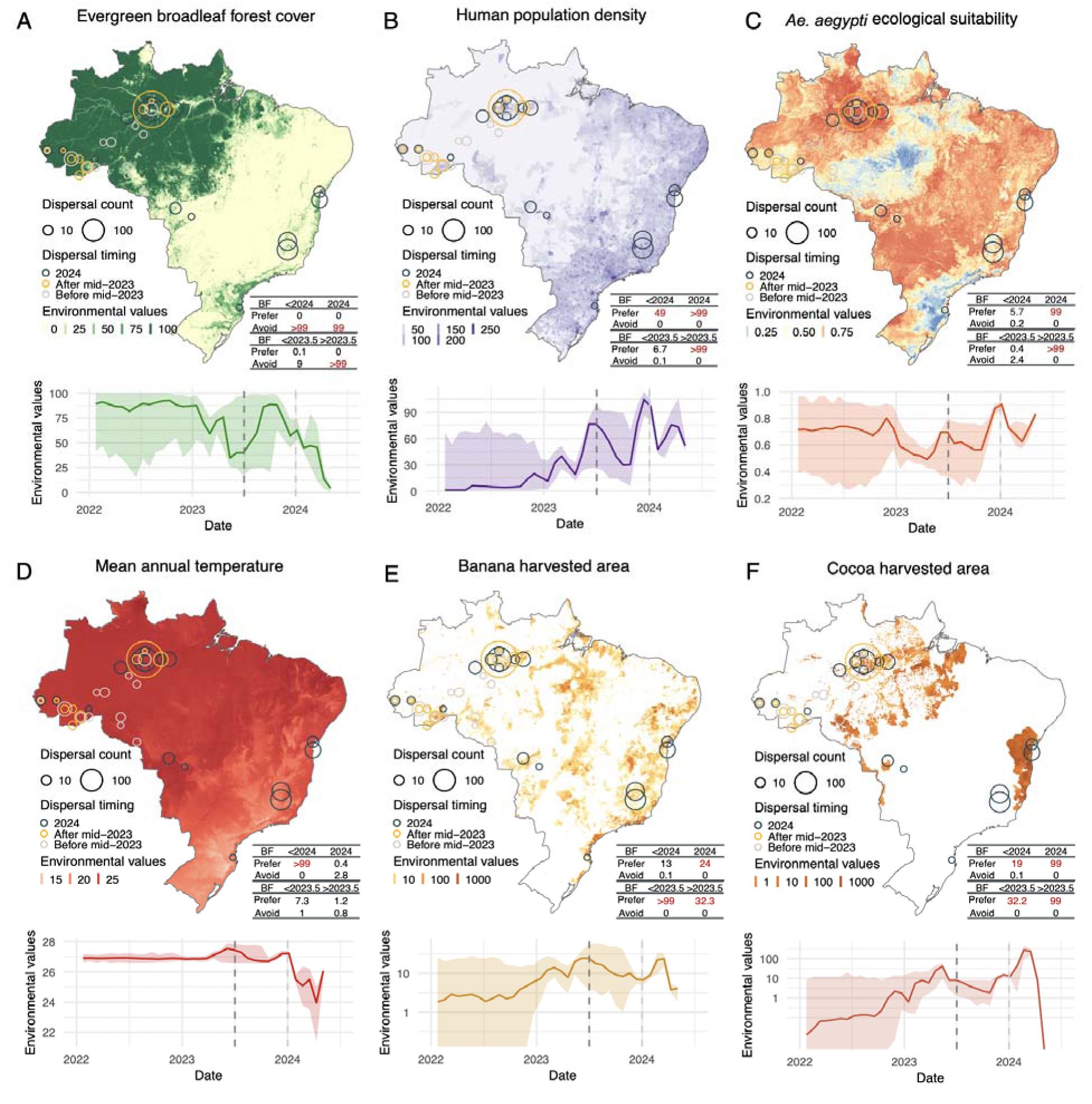
Environmental conditions associated with OROV lineage dispersal locations over time (<u>fo segment S</u>). Figure panels show the spatial distribution of six main environmental factors (units specified): evergreen broadleaf forest cover (%) **(A)**, human population density (normalised between 0 and 255 per km^2^ for visual clarity) (**B**), *Ae. aegypti* ecological suitability (probability of occurrence) (**C**), mean annual temperature (°C) (**D**), banana harvested area (hectares - log) (**E**), and cocoa harvested area (hectares - log) (**F**) in the top rows. Circles on the map depict the end node of dispersal locations inferred by continuous phylogeography, sized by the number of dispersal events in an area, and coloured by the timing of the event. Bottom rows of each figure panel are line graphs depicting the environmental covariates associated with the locations of OROV lineage dispersal events i Brazil. Each plot illustrates how specific ecological conditions have changed over time (2022-2024) at the sites of viral lineage dispersal. The embedded tables show the association between environmental conditions and the dispersal location of inferred OROV lineages. Based on the analysis of 100 posterior trees obtained from continuous phylogeographic inference, the table reports Bayes factor (BF) supports for association between environmental raster values and tree node locations. Following the scale of interpretation of Kass and Raftery (*24*), we highlight BF values >20 considered as strong supports.

**Figure S6.**
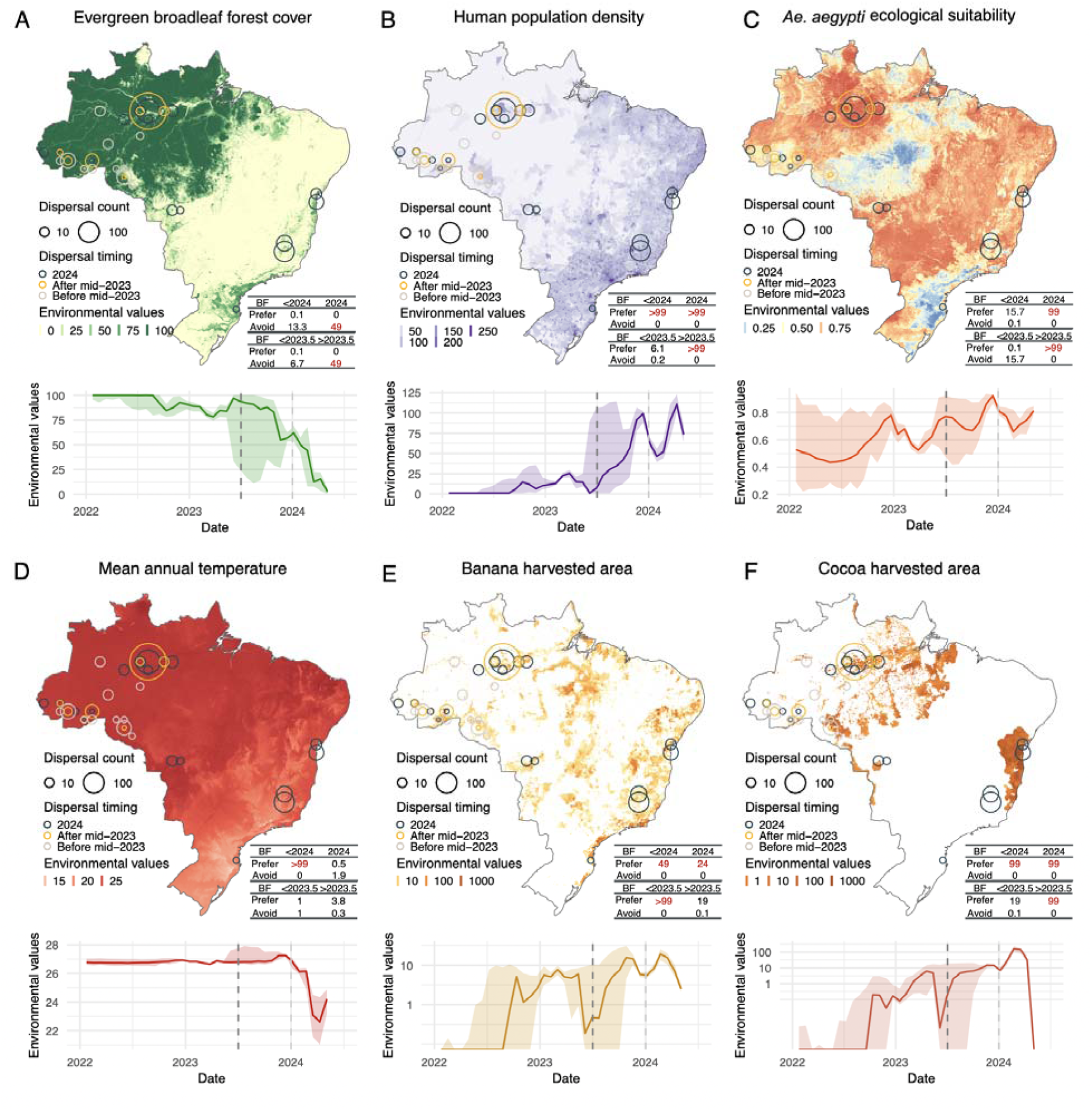
Environmental conditions associated with OROV lineage dispersal locations over time (<u>fo segment L</u>). Figure panels show the spatial distribution of six main environmental factors (units specified): evergreen broadleaf forest cover (%) **(A)**, human population density (normalised between 0 and 255 per km^2^ for visual clarity) (**B**), *Ae. aegypti* ecological suitability (probability of occurrence) (**C**), mean annual temperature (°C) (**D**), banana harvested area (hectares - log) (**E**), and cocoa harvested area (hectares - log) (**F**) in the top rows. Circles on the map depict the end node of dispersal locations inferred by continuous phylogeography, sized by the number of dispersal events in an area, and coloured by the timing of the event. Bottom rows of each figure panel are line graphs depicting the environmental covariates associated with the locations of OROV lineage dispersal events in Brazil. Each plot illustrates how specific ecological conditions have changed over time (2022-2024) at the sites of viral lineage dispersal. The embedded tables show the association between environmental conditions and the dispersal location of inferred OROV lineages. Based on the analysis of 100 posterior trees obtained from continuous phylogeographic inference, the table reports Bayes factor (BF) supports for association between environmental raster values and tree node locations. Following the scale of interpretation of Kass and Raftery (*24*), we highlight BF values >20 considered as strong supports.

**Figure S7.**
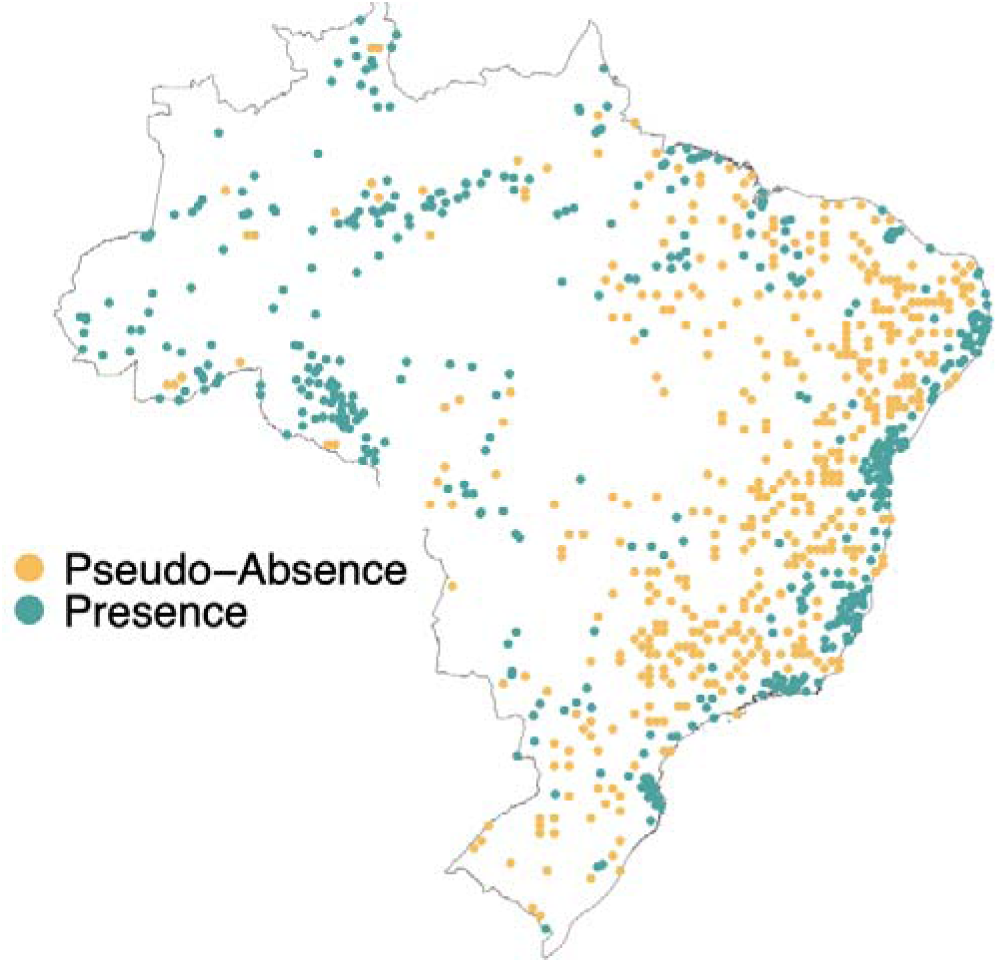
Distribution of disease presence and pseudo-absence points. We generated pseudo-absence points at a 1:1 ratio with presence points by sampling from the distribution of presence points and th kernel density estimate of human population density.

**Figure S8.**
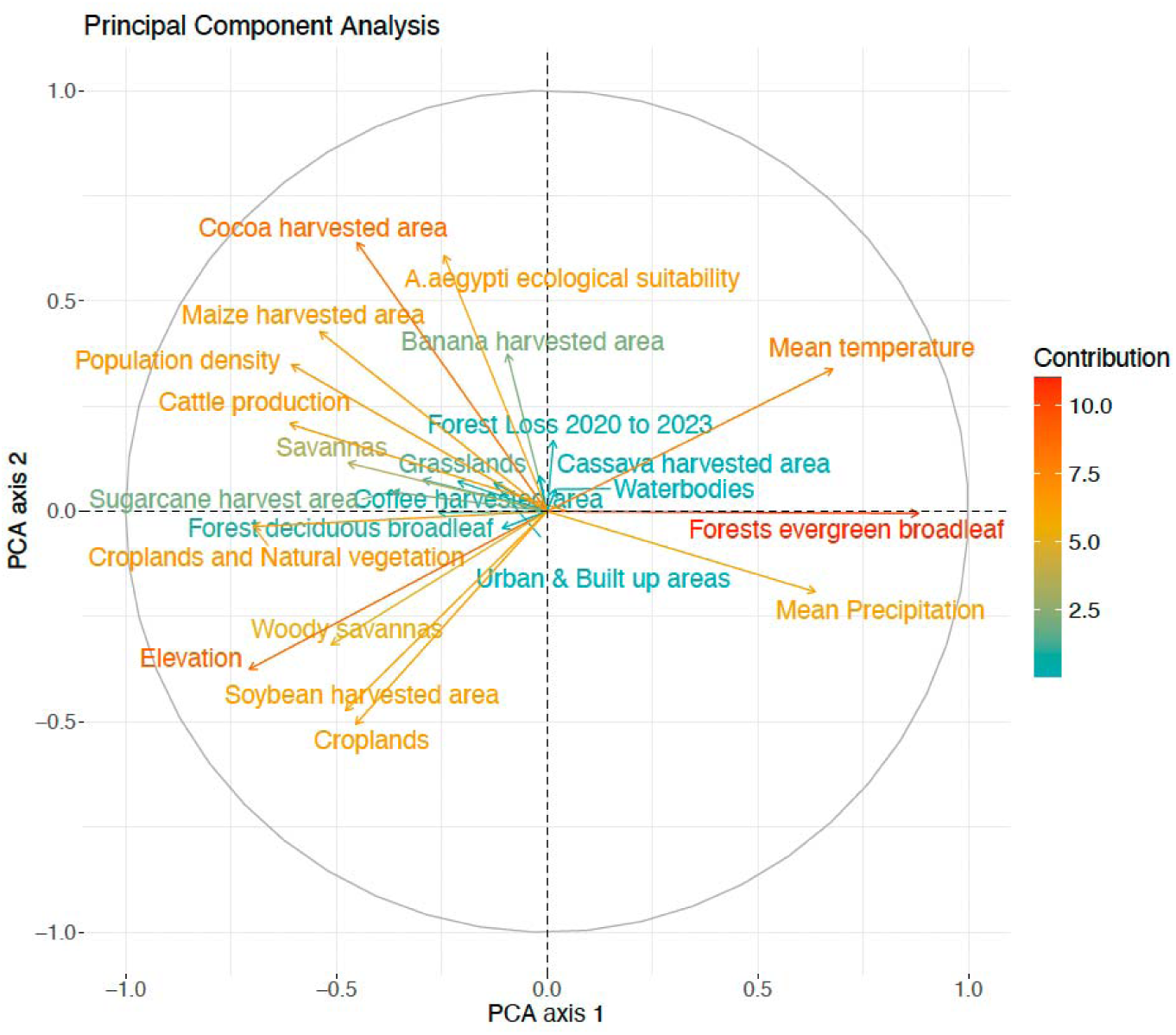
Principal Component Analysis (PCA) plot illustrating the relationships between variables. Arrows that lie within the same quadrant or are positioned close to each other indicate a higher correlation among the corresponding variables. Furthermore, longer arrows signify a greater contribution of those variables to the principal components, highlighting their discrimination in the overall dataset.

**Figure S9.**
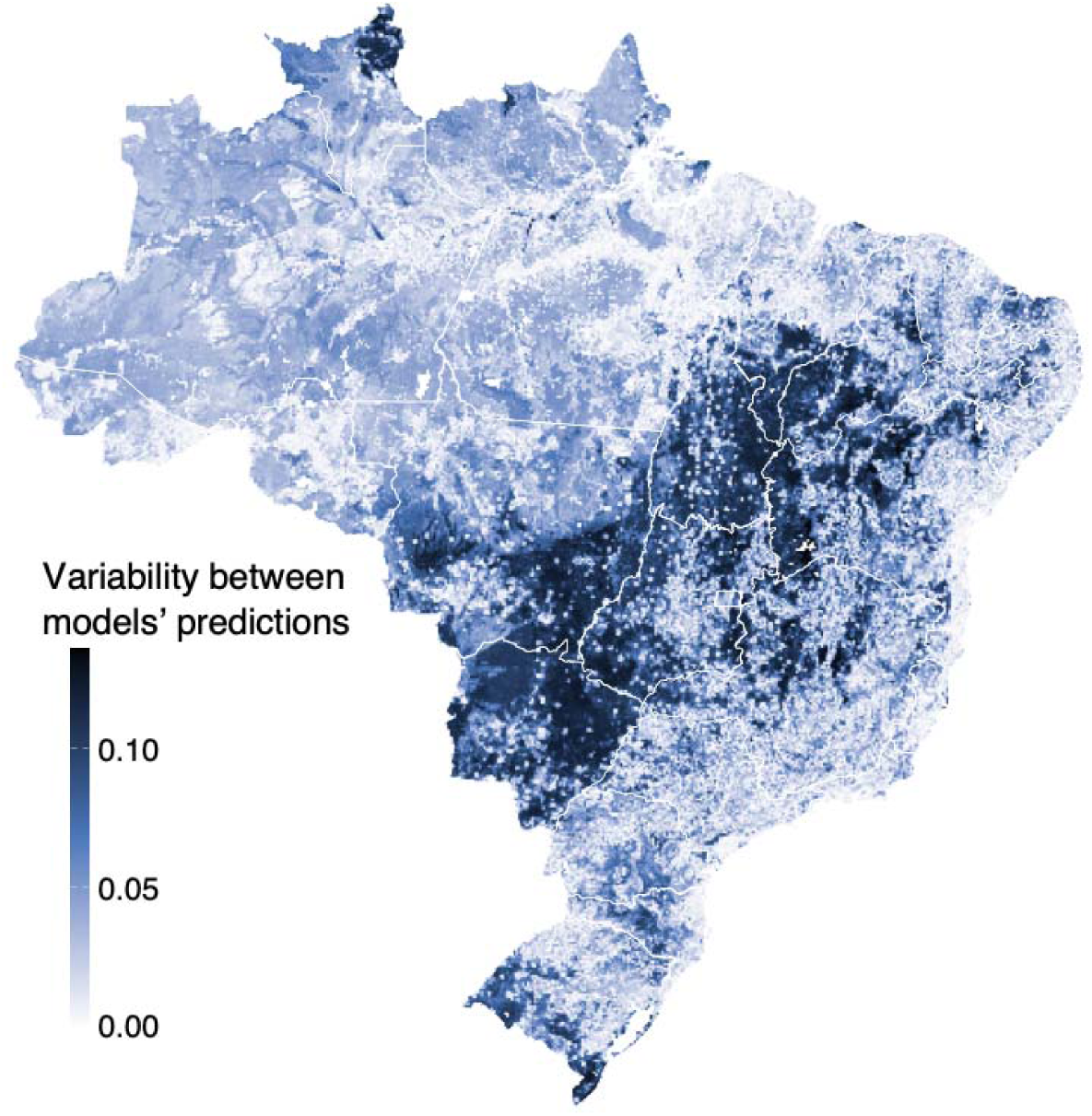
Ecological niche models variability. The degree of variability in suitability prediction values among the models in our ensemble, highlighting areas where different models either converge or diverge in their predictions.

**Figure S10.**
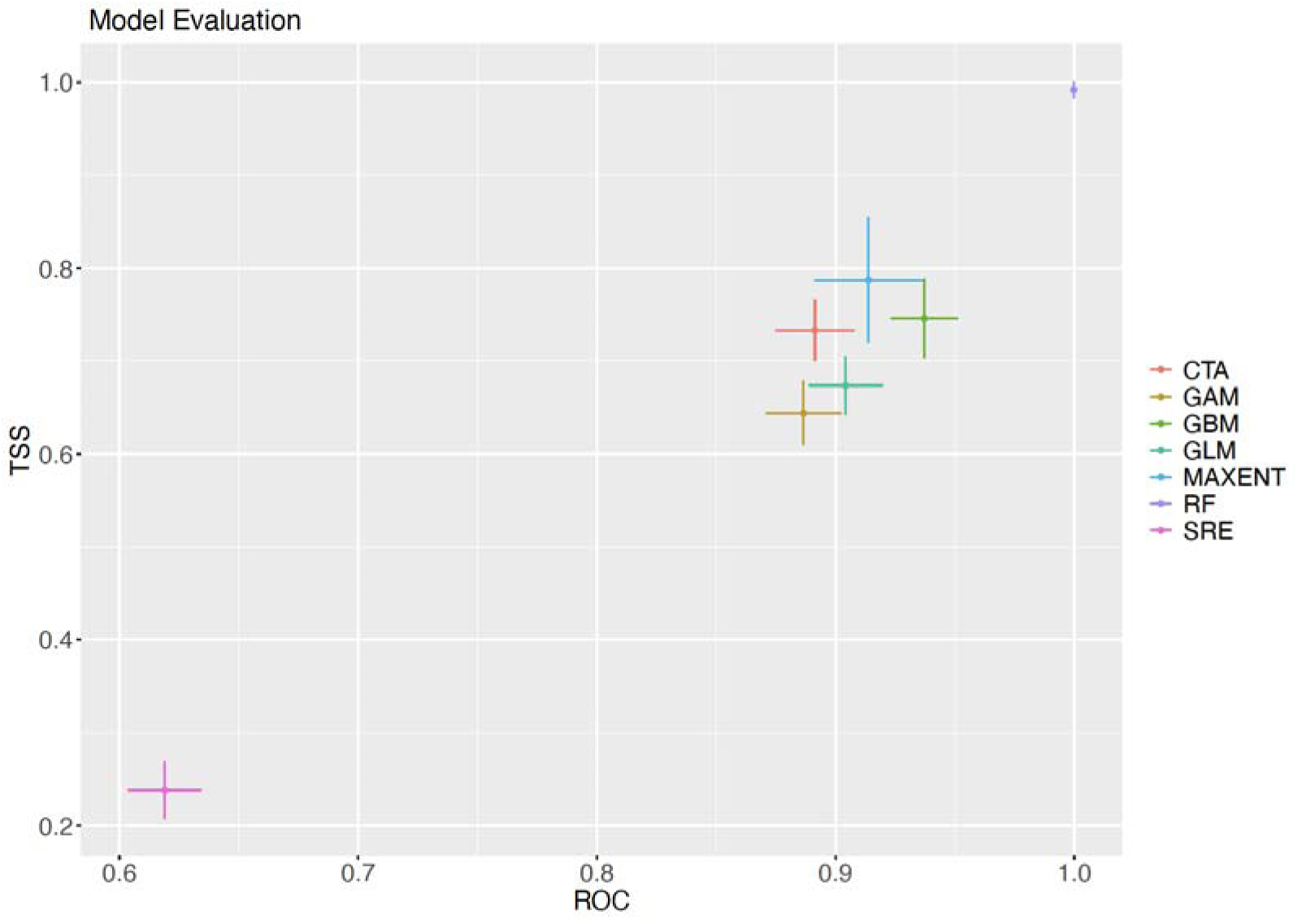
Ecological niche models evaluation results. Results from block cross-validation of the individual environmental niche models of the full model (using all data points). The x- and y-axis show the True Skill Statistic (TSS) and area under the Receiver Operating Characteristic (ROC) curve, respectively.

**Supplementary Table S1:**
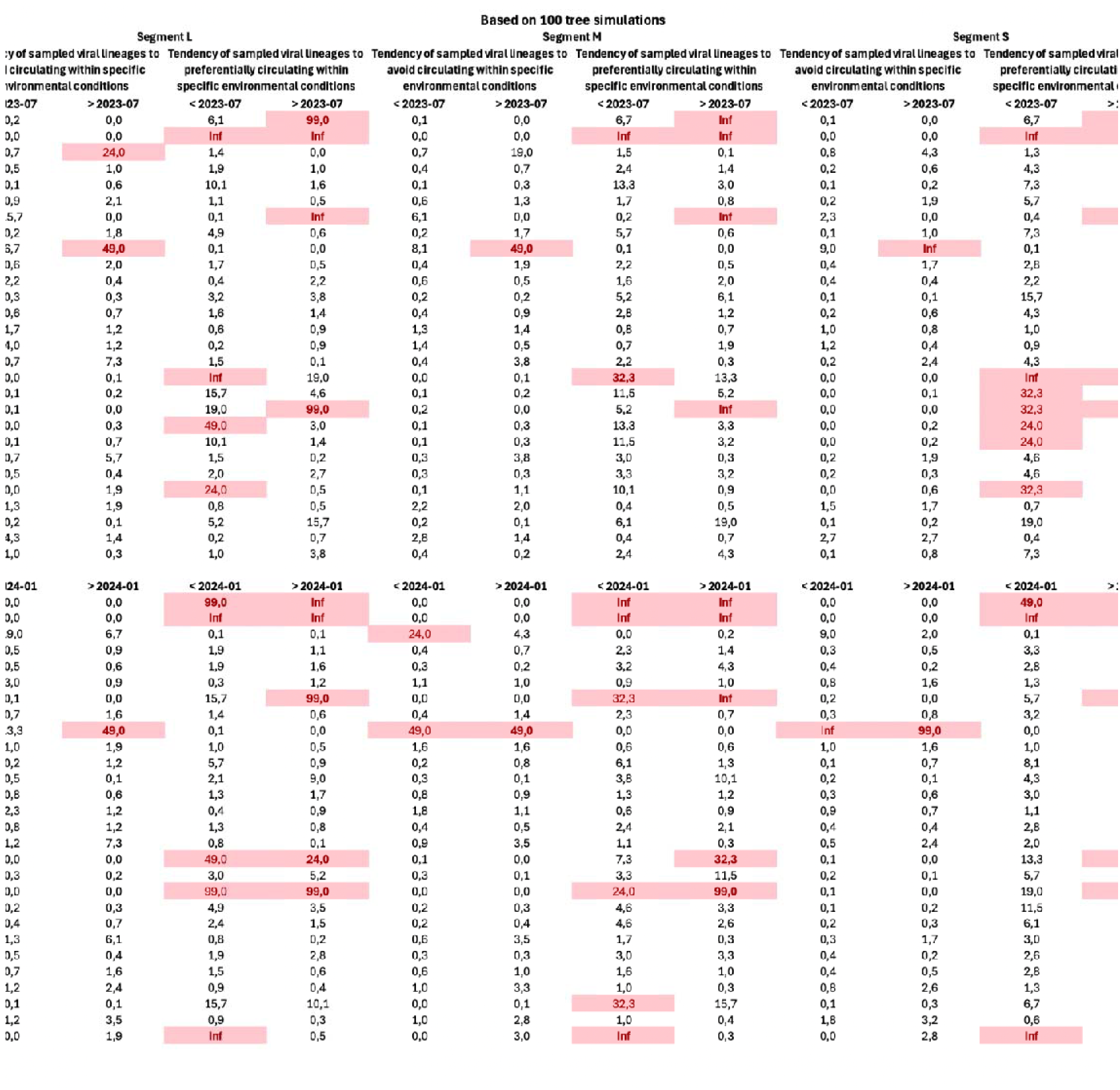

**Supplementary Table S2:**
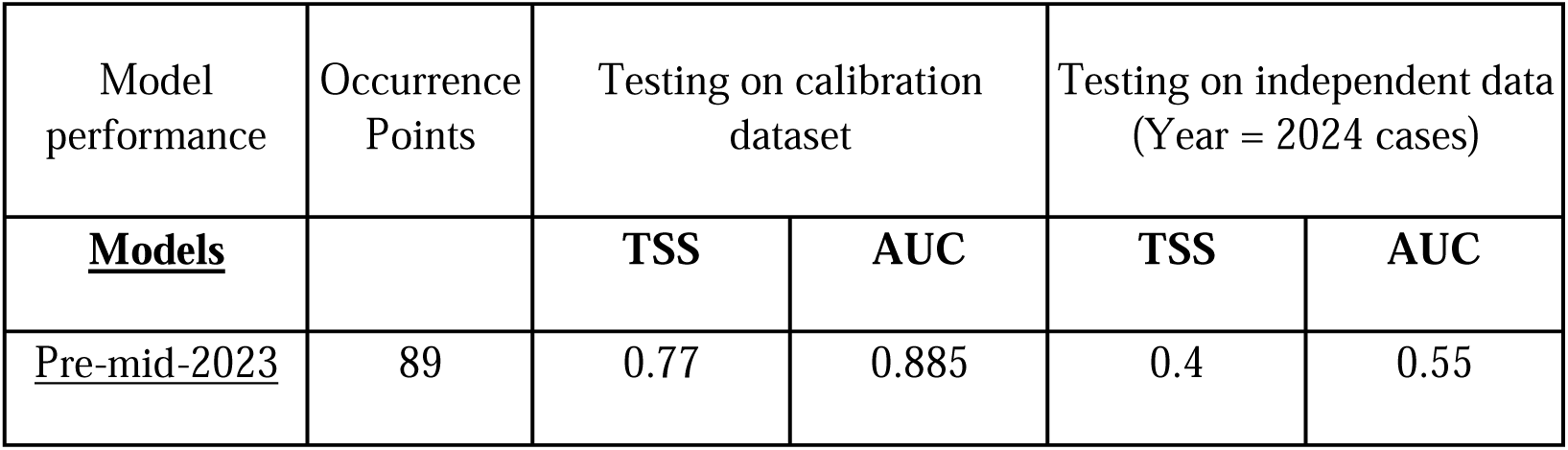
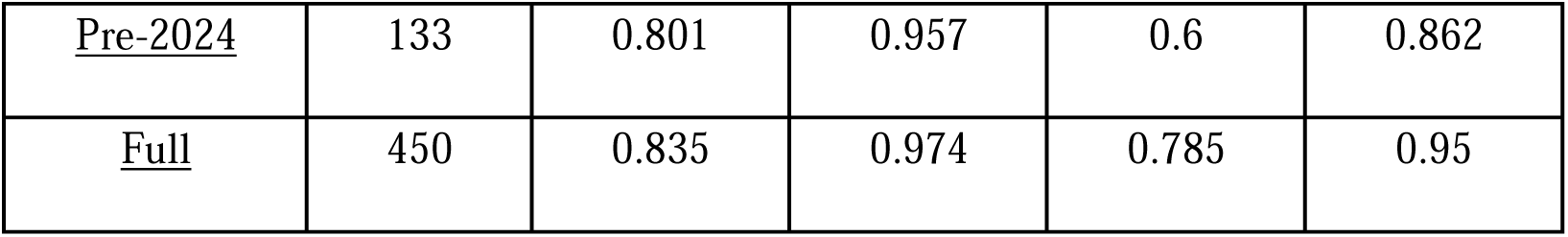
Model evaluation metrics (TSS and ROC) for the ensemble models, with results from testing on both the training/calibration datasets (left) and an independent dataset from 2024 cases (right). The models were evaluated in three stages: points collected before mid-2023, points collected before 2024, and all available points combined.

**Supplementary Table S3:**
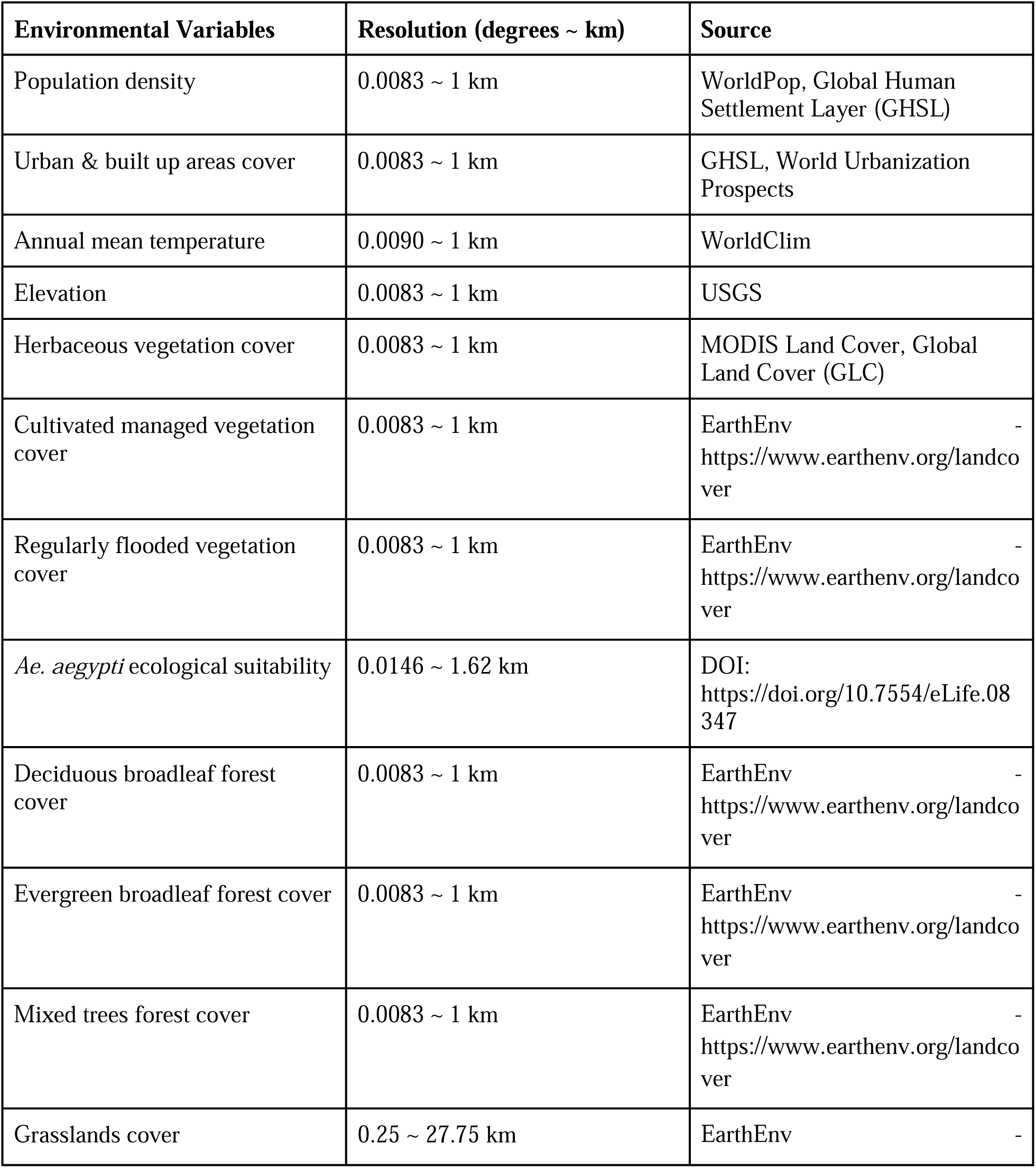
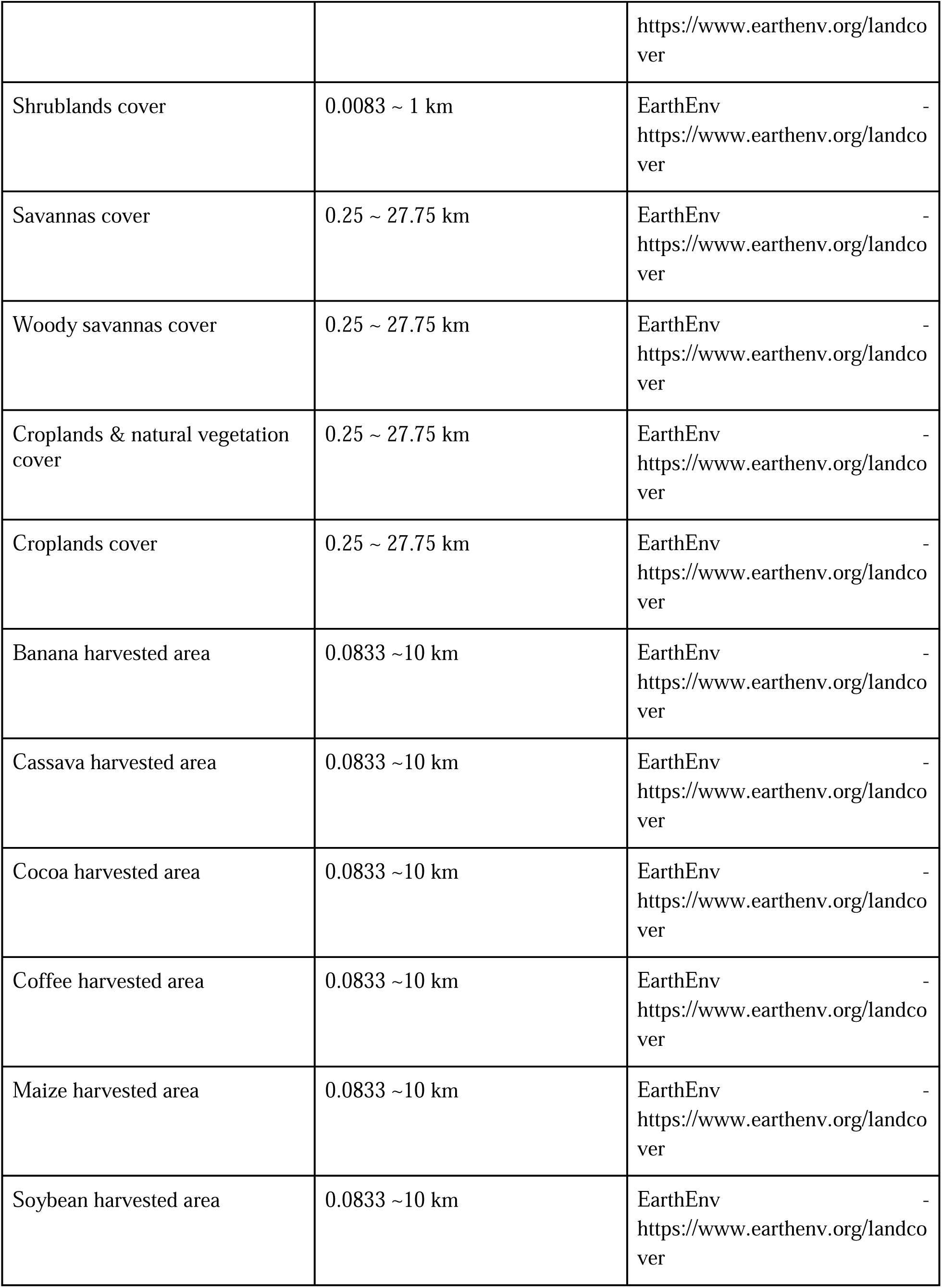
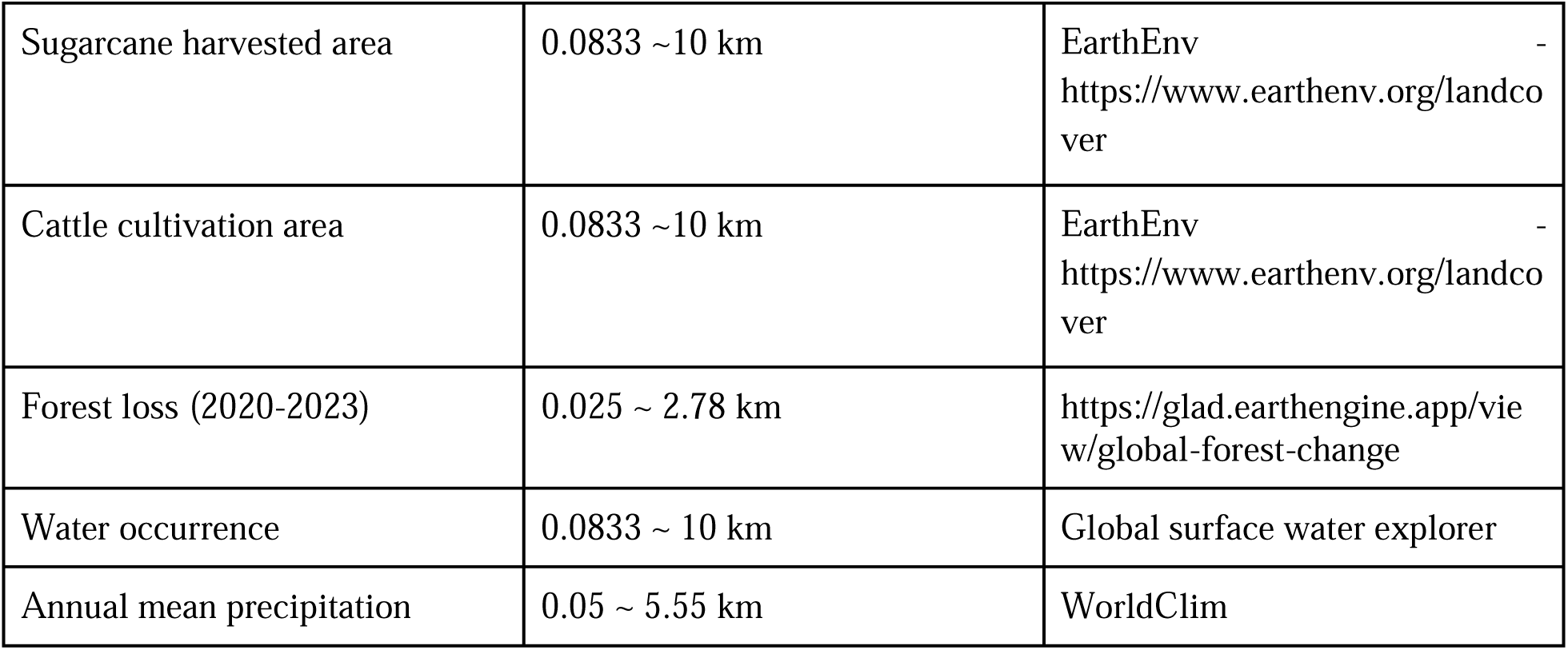
Environmental variables used in the study, with respective resolutions and data sources.

## CLIMADE Consortium Contributing Authors

Luiz C J Alcantara^5,6^, Marta Giovanetti^5,6^, Edward C Holmes^6^, Vagner Fonseca^7^, Tanya Golubchi^6^, Samuel Oyola^8^,, Jenicca Poongavanan^1^, Graeme Dor^1^, Gaspary Mwanyika^1^, José Lourenco^10^, Frank Tanser^1^, Richard Lessells^2^, Abdou Padane^11,^ Ambroise Ahouidi^11^, Abdualmoniem O A Musa^12^, Adugna Abera^13^, Allan Campbell^14^, Aloysious S Semaganda^15^, Argentina F Muianga^16^, Bernard Onoja^17^, Birhanu D Alemu^18^, Darren Martin^19^, Mohamed Z Alimohamed^20^, Fredy B N Simo^21^, Girma Godebo^22^, James Ayei Maror^23,^ John Oludele^24^, Joseph Fokam^25^, Kenneth K Maeka^26^, Lavanya Singh^2^, Martin Faye^27^, Michael Owusu^28^, Michel N Dikongo^29^, Molalegne Bitew ^30^, Nkuurunziza Jerome^31^, Nokuzola Mbhele^19^, Oyewale Tomori^32^, Ramuth Magalutcheemee^33^, Sara A Abuelmaali^34^, Wolfgang Preiser^35^

^1^ Centre for Epidemic Response Innovation (CERI), School for Data Science and Computational Thinking, Stellenbosch University, South Africa;

^2^ KwaZulu-Natal Research Innovation and Sequencing Platform (KRISP), University of KwaZulu-Natal, South Africa.

^3^ Duke Human Vaccine Institute, Duke University, Durham, NC 27710, USA

^4^ Institute of Social and Preventive Medicine (ISPM), University in Bern, Switzerland.

^5^ Laboratório de Flavivírus, Instituto Oswaldo Cruz, Fundação Oswaldo Cruz, Rio de Janeiro, Brazil

^6^ Instituto Rene Rachou, Fundação Oswaldo Cruz, Belo Horizonte, Minas Gerais, Brazil.

^7^ Marie Bashir Institute for Infectious Diseases and Biosecurity, School of Life and Environmental Sciences and School of Medical Sciences, University of Sydney, Sydney, NSW, Australia

^8^ Organização Pan-Americana da Saúde/Organização Mundial da Saúde, Brasília, Distrito Federal, Brazil.8. Organização Pan-Americana da Saúde/Organização Mundial da Saúde, Brasília, Distrito Federal, Brazil.

^9^ International Livestock Research Institute (ILRI), Kenya

^10^ CBR (Biomedical Research Centre), Universidade Católica Portuguesa, Oeiras, Portugal.

^11^ Institute de Recherche en Santé, de Surveillance Épidémiologique et de Formations (IRESSEF), Senegal.

^12^ General Administration of Laboratories and Blood Banks, Ministry of Health, Kassala state, Sudan.

^13^ Ethiopian Public Health Institute, Ethiopia.

^14^ Central Public Health Reference Laboratory, Sierra Leone

^15^ National Health Laboratories and Diagnostic Services - Central Public Health Laboratories, Uganda

^16^ Instituto Nacional de Saude, Mozambique), Aziza John Samson, Tanzania

^17^ University of Ibadan, Nigeria.

^18^ PATH, Ethiopia.

^19^ University of Cape Town, South Africa.

^20^ Muhimbili University of Health and Allied Sciences, Tanzania.

^21^ Centre for Research in Infectious Disease, Cameroon.

^22^ Wachemo University, Ethiopia.

^23^ National Public Health Laboratory, South Sudan.

^24^ Instituto Nacional de Saude, Mozambique.

^25^ Chantal BIYA International Reference Centre (CIRCB), Cameroon.

^26^ National Microbiology Reference Laboratory, Ministry of Health, Zimbabwe.

^27^ Institute Pasteur de Dakar, Senegal.

^28^ Kwame Nkrumah University of Science and Technology, Ghana.

^29^ Biologiste, Gabon.

^30^ Bio and Emerging Technology Institute, Ethiopia.

^31^ Hope Africa University, National Institute of Public Health Reference Laboratory, Burundi.

^32^ African Centre of Excellence for Genomics of Infectious (ACEGID) Redeemer’s University, Nigeria.

^33^ Ministry of Health, Mauritius.

^34^ National Public Health Laboratory, Sudan.

^35^ University of Stellenbosch / National Health Laboratory Service, Tygerberg, South Africa.

## References

1. J. F. Travassos da Rosa, W. M. de Souza, F. de P. Pinheiro, M. L. Figueiredo, J. F. Cardoso, G. O. Acrani, M. R. T. Nunes, Oropouche virus: clinical, epidemiological, and molecular aspects of a neglected orthobunyavirus. Am. J. Trop. Med. Hyg. 96, 1019–1030 (2017).

2. About Oropouche | Oropouche | CDC, (available at https://www.cdc.gov/oropouche/about/index.html).

3. H. M. Moreira, G. Sgorlon, J. A. S. Queiroz, T. P. Roca, J. Ribeiro, K. S. Teixeira, A. M. Passos-Silva, A. Araújo, N. W. F. Gasparelo, A. de O. Dos Santos, C. A. B. Lugtenburg, R. A. Roque, J. M. Villalobos Salcedo, D. B. Pereira, D. Vieira, Outbreak of Oropouche virus in frontier regions in western Amazon. Microbiol. Spectr. 12, e0162923 (2024).

4. H. Sakkas, P. Bozidis, A. Franks, C. Papadopoulou, Oropouche fever: A review. Viruses. 10 (2018), doi:10.3390/v10040175.

5. B. F. Cardoso, O. P. Serra, L. B. da S. Heinen, N. Zuchi, V. C. de Souza, F. G. Naveca, M. A. M. dos Santos, R. D. Slhessarenko, Detection of Oropouche virus segment S in patients and inCulex quinquefasciatus in the state of Mato Grosso, Brazil. Mem. Inst. Oswaldo Cruz. 110, 745–754 (2015).

6. Oropouche virus disease - Region of the Americas, (available at https://www.who.int/emergencies/disease-outbreak-news/item/2024-DON530).

7. J. Usuga, D. Limonta, L. S. Perez-Restrepo, K. A. Ciuoderis, I. Moreno, A. Arevalo, V. Vargas, M. G. Berg, G. A. Cloherty, J. P. Hernandez-Ortiz, J. E. Osorio, Co-Circulation of 2 Oropouche Virus Lineages, Amazon Basin, Colombia, 2024. Emerging Infect. Dis. 30 (2024), doi:10.3201/eid3011.240405.

8. A. C. Bandeira, A. C. F. N. da S. Barbosa, M. Souza, R. da C. Saavedra, F. M. Pereira, S. P. de O. Santos, A. L. e S. de Mello, S. M. O. da Purificação, D. R. de Souza, A. A. de A. Lessa, N. R. Guimarães, V. Fonseca, M. Giovanetti, L. C. J. Alcantara, L. M. R. Tome, F. C. de M. Iani, R. M. Barros, R. R. Fonseca, J. P. de Jesus, M. L. V. Araújo, Clinical profile of Oropouche Fever in Bahia, Brazil: unexpected fatal cases (2024), doi:10.1590/SciELOPreprints.9342.

9. F. E. das Neves Martins, J. O. Chiang, B. T. D. Nunes, B. de F. R. Ribeiro, L. C. Martins, L. M. N. Casseb, D. F. Henriques, C. S. de Oliveira, E. L. N. Maciel, R. da S. Azevedo, L. de C. C. Cravo, A. R. F. Barreto, A. L. S. Pessoa, A. J. M. Filho, J. R. de Sousa, L. Schuler-Faccini, J. A. S. Quaresma, P. F. da Costa Vasconcelos, R. do S. da Silva Azevedo, Newborns with microcephaly in Brazil and potential vertical transmission of Oropouche virus: a case series. Lancet Infect. Dis. (2024), doi:10.1016/S1473-3099(24)00617-0.

10. F. C. de M. Iani, F. Mota Pereira, E. C. de Oliveira, J. T. Nascimento Rodrigues, M. Hoffmann Machado, V. Fonseca, T. E. Ribeiro Adelino, N. Rocha Guimaraes, L. M. Ribeiro Tome, M. K. Astete Gomez, V. Brandao Nardy, A. A. Ribeiro, A. Rosewell, A. G. A. Ferreira, A. Leal e Silva de Mello, B. Machado Moura Fernandes, C. F. C. de Albuquerque, D. dos Santos Pereira, E. Carvalho Pimentel, F. G. Mesquita Lima, M. Giovanetti, Rapid spatial Expansion Beyond the Amazon Basin: Oropouche Virus joins other main arboviruses in epidemic activity across the Americas. medRxiv (2024), doi:10.1101/2024.08.02.24311415.

11. F. G. Naveca, T. A. P. de Almeida, V. Souza, V. Nascimento, D. Silva, F. Nascimento, M. Mejía, Y. S. de Oliveira, L. Rocha, N. Xavier, J. Lopes, R. Maito, C. Meneses, T. Amorim, L. Fé, F. S. Camelo, S. C. de A. Silva, A. X. de Melo, L. G. Fernandes, M. A. A. de Oliveira, G. Bello, Human outbreaks of a novel reassortant Oropouche virus in the Brazilian Amazon region. Nat. Med. (2024), doi:10.1038/s41591-024-03300-3.

12. M. Giovanetti, F. Pinotti, C. Zanluca, V. Fonseca, T. Nakase, A. C. Koishi, M. Tscha, G. Soares, G. G. Dorl, A. E. M. L. Marques, R. Sousa, T. E. R. Adelino, J. Xavier, C. de Oliveira, S. Patroca, N. R. Guimaraes, H. Fritsch, M. A. Mares-Guia, F. Levy, P. H. Passos, C. N. Duarte Dos Santos, Genomic epidemiology unveils the dynamics and spatial corridor behind the Yellow Fever virus outbreak in Southern Brazil. Sci. Adv. 9, eadg9204 (2023).

13. M. A. Files, C. A. Hansen, V. C. Herrera, C. Schindewolf, A. D. T. Barrett, D. W. C. Beasley, N. Bourne, G. N. Milligan, Baseline mapping of Oropouche virology, epidemiology, therapeutics, and vaccine research and development. npj Vaccines. 7, 38 (2022).

14. J. L.-H. Tsui, R. E. Pena, M. Moir, R. P. D. Inward, E. Wilkinson, J. E. San, J. Poongavanan, S. Bajaj, B. Gutierrez, A. Dasgupta, T. de Oliveira, M. U. G. Kraemer, H. Tegally, P. Sambaturu, Impacts of climate change-related human migration on infectious diseases. Nat. Clim. Chang. 14, 793–802 (2024).

15. K. M. Wesselmann, I. Postigo-Hidalgo, L. Pezzi, E. F. de Oliveira-Filho, C. Fischer, X. de Lamballerie, J. F. Drexler, Emergence of Oropouche fever in Latin America: a narrative review. Lancet Infect. Dis. 24, e439–e452 (2024).

16. C. E. S. Walsh, M. A. Robert, R. C. Christofferson, Observational Characterization of the Ecological and Environmental Features Associated with the Presence of Oropouche Virus and the Primary Vector Culicoides paraenesis: Data Synthesis and Systematic Review. Trop. Med. Infect. Dis. 6 (2021), doi:10.3390/tropicalmed6030143.

17. D. Romero-Alvarez, L. E. Escobar, Emergent viruses in America: The case of Oropouche virus. Int. J. Infect. Dis. 73, 98 (2018).

18. C. A. V. Aybar, M. J. D. Juri, M. S. L. de Grosso, G. R. Spinelli, Spatial and Temporal Distribution of *Culicoides insignis* and *Culicoides paraensis* in the Subtropical Mountain Forest of Tucumán, Northwestern Argentina. Florida Entomologist. 94, 1018–1025 (2011).

19. M. E. Gorris, A. W. Bartlow, S. D. Temple, D. Romero-Alvarez, D. P. Shutt, J. M. Fair, K. A. Kaufeld, S. Y. Del Valle, C. A. Manore, Updated distribution maps of predominant Culex mosquitoes across the Americas. Parasit. Vectors. 14, 547 (2021).

20. S. Dellicour, P. Bastide, P. Rocu, D. Fargette, O. J. Hardy, M. A. Suchard, S. Guindon, P. Lemey, How fast are viruses spreading in the wild? BioRxiv (2024), doi:10.1101/2024.04.10.588821.

21. E. N. Gallichotte, G. Ebel, C. J. Carlson, Vector competence for Oropouche virus: a systematic review of pre-2024 experiments. medRxiv (2024), doi:10.1101/2024.10.17.24315699.

22. S. F. de Mendonça, M. N. Rocha, F. V. Ferreira, T. H. J. F. Leite, S. C. G. Amadou, P. H. F. Sucupira, J. T. Marques, A. G. A. Ferreira, L. A. Moreira, Evaluation of Aedes aegypti, Aedes albopictus, and Culex quinquefasciatus Mosquitoes Competence to Oropouche virus Infection. Viruses. 13 (2021), doi:10.3390/v13050755.

23. P. Rozo-Lopez, Y. Park, B. S. Drolet, Effect of Constant Temperatures on Culicoides sonorensis Midge Physiology and Vesicular Stomatitis Virus Infection. Insects. 13 (2022), doi:10.3390/insects13040372.

24. R. E. Kass, A. E. Raftery, Bayes Factors. J. Am. Stat. Assoc. 90, 773–795 (1995).

25. S. Dellicour, C. Troupin, F. Jahanbakhsh, A. Salama, S. Massoudi, M. K. Moghaddam, G. Baele, P. Lemey, A. Gholami, H. Bourhy, Using phylogeographic approaches to analyse the dispersal history, velocity and direction of viral lineages - Application to rabies virus spread in Iran. Mol. Ecol. 28, 4335–4350 (2019).

26. A. L. Hoch, D. R. Roberts, F. P. Pinheiro, Breeding sites of Culicoides paraensis and options for control by environmental management. Bulletin of the Pan American … (1986).

27. S. Carpenter, M. H. Groschup, C. Garros, M. L. Felippe-Bauer, B. V. Purse, Culicoides biting midges, arboviruses and public health in Europe. Antiviral Res. 100, 102–113 (2013).

28. A. L. Hoch, D. R. Roberts, Host-seeking behavior and seasonal abundance of Culicoides paraensis(Diptera: Ceratopogonidae) in Brazil. Journal of the American … (1990).

29. G. C. Scachetti, J. Forato, I. M. Claro, X. Hua, B. B. Salgado, A. Vieira, C. L. Simeoni, A. R. C. Barbosa, I. L. Rosa, G. F. de Souza, L. C. N. Fernandes, A. C. H. de Sena, S. C. Oliveira, C. M. L. Singh, S. T. S. de Lima, R. de Jesus, M. A. Costa, R. B. Kato, J. F. Rocha, L. C. Santos, W. M. de Souza, Re-emergence of Oropouche virus between 2023 and 2024 in Brazil: an observational epidemiological study. Lancet Infect. Dis. (2024), doi:10.1016/S1473-3099(24)00619-4.

30. Why did an obscure virus explode in Latin America? New study offers clues | Science | AAAS, (available at https://www.science.org/content/article/why-did-obscure-virus-explode-latin-america-new-study-offers-clues).

31. D. Romero-Alvarez, L. E. Escobar, A. J. Auguste, S. Y. Del Valle, C. A. Manore, Transmission risk of Oropouche fever across the Americas. Infect. Dis. Poverty. 12, 47 (2023).

32. B. Gutierrez, E. L. Wise, S. T. Pullan, C. H. Logue, T. A. Bowden, M. Escalera-Zamudio, G. Trueba, M. R. T. Nunes, N. R. Faria, O. G. Pybus, Evolutionary dynamics of oropouche virus in south america. J. Virol. 94 (2020), doi:10.1128/JVI.01127-19.

33. D. Romero-Alvarez, L. E. Escobar, Vegetation loss and the 2016 Oropouche fever outbreak in Peru. Mem. Inst. Oswaldo Cruz. 112, 292–298 (2017).

34. M. C. Castro, A. S. Lima Neto, Unprecedented spread and genetic evolution of the Oropouche virus. Nat. Med. (2024), doi:10.1038/s41591-024-03336-5.

35. T. Gräf, E. Delatorre, C. do N. Ferreira, A. Rossi, B. R. Pizzato, V. Nascimento, V. Souza, G. B. de Lima, F. Z. Dezordi, A. F. da Silva, C. N. L. de Morais, I. Arantes, M. H. Machado, D. B. Rovaris, M. M. Presibella, N. F. Q. Marques, E. G. Pouzato, J. Stadinicki, R. Ribeiro-Rodrigues, T. de J. Souza, F. G. Naveca, Long-Range Spread and Sustained Transmission of Oropouche Virus Outside the Endemic Brazilian Amazon Region (2024), doi:10.2139/ssrn.4932381.

## Supplementary References

36. K. Katoh, K. Misawa, K. Kuma, T. Miyata, MAFFT: A novel method for rapid multiple sequence alignment based on fast Fourier transform. Nucleic Acids Res. 30, 3059–3066 (2002).

37. K. Katoh, D. M. Standley, MAFFT multiple sequence alignment software version 7: improvements in performance and usability. Mol. Biol. Evol. 30, 772–780 (2013).

38. A. Larsson, AliView: a fast and lightweight alignment viewer and editor for large datasets. Bioinformatics. 30, 3276–3278 (2014).

39. Brazil’s Air Travel Hits Pre-Pandemic Heights - The Rio Times, (available at https://www.riotimesonline.com/brazil-news/brazils-air-travel-hits-pre-pandemic-heights/).

40. M. A. Suchard, P. Lemey, G. Baele, D. L. Ayres, A. J. Drummond, A. Rambaut, Bayesian phylogenetic and phylodynamic data integration using BEAST 1.10. Virus Evol. 4, vey016 (2018).

41. A. Rambaut, T. T. Lam, L. Max Carvalho, O. G. Pybus, Exploring the temporal structure of heterochronous sequences using TempEst (formerly Path-O-Gen). Virus Evol. 2, vew007 (2016).

42. P. Lemey, A. Rambaut, J. J. Welch, M. A. Suchard, Phylogeography takes a relaxed random walk in continuous space and time. Mol. Biol. Evol. 27, 1877–1885 (2010).

43. S. Dellicour, M. S. Gill, N. R. Faria, A. Rambaut, O. G. Pybus, M. A. Suchard, P. Lemey, Relax, Keep Walking - A Practical Guide to Continuous Phylogeographic Inference with BEAST. Mol. Biol. Evol. 38, 3486–3493 (2021).

44. A. Rambaut, A. J. Drummond, D. Xie, G. Baele, M. A. Suchard, Posterior summarization in Bayesian phylogenetics using Tracer 1.7. Syst. Biol. 67, 901–904 (2018).

45. S. Dellicour, R. Rose, N. R. Faria, P. Lemey, O. G. Pybus, SERAPHIM: studying environmental rasters and phylogenetically informed movements. Bioinformatics. 32, 3204–3206 (2016).

46. N. S. Trovão, M. A. Suchard, G. Baele, M. Gilbert, P. Lemey, Bayesian Inference Reveals Host-Specific Contributions to the Epidemic Expansion of Influenza A H5N1. Mol. Biol. Evol. 32, 3264–3275 (2015).

47. C. Merow, M. J. Smith, J. A. Silander, A practical guide to MaxEnt for modeling species’ distributions: what it does, and why inputs and settings matter. Ecography. 36, 1058–1069 (2013).

48. A. Guisan, W. Thuiller, N. E. Zimmermann, Habitat suitability and distribution models: with applications in R (Cambridge University Press, Cambridge, 2017), *Ecology, Biodiversity and Conservation*.

49. M. Marmion, M. Parviainen, M. Luoto, R. K. Heikkinen, W. Thuiller, Evaluation of consensus methods in predictive species distribution modelling. Diversity and Distributions. 15, 59–69 (2009).

50. R. Muscarella, P. J. Galante, M. Soley-Guardia, R. A. Boria, J. M. Kass, M. Uriarte, R. P. Anderson, ENMeval: An R package for conducting spatially independent evaluations and estimating optimal model complexity for Maxent ecological niche models. Methods Ecol. Evol. 5, 1198–1205 (2014).

51. B. Gregorutti, B. Michel, P. Saint-Pierre, Correlation and variable importance in random forests. Stat. Comput. 27, 659–678 (2017).

