## Supplementary Information for "Dynamics and ecology of a multi-stage expansion of Oropouche virus in Brazil"

**Supplementary Materials**

Materials and Methods

Supplementary Figures S1 to S10

Supplementary Tables S1 to S3

References (35–51)

CLIMADE Consortium Contributing Authors

**Materials and Methods**

**Oropouche Virus genomic data**

Complete genome sequences of the S, M, and L segments of the Oropouche virus (OROV), obtained from the first extra-Amazon OROV cases reported in the states of Bahia (Northeast Brazil), Minas Gerais (Southeast Brazil), Mato Grosso (Midwest Brazil), and Paraná (South Brazil), were combined with the corresponding segments of recently published full-length OROV sequences belonging to the Brazilian 2022-2024 sublineage [(*10*)](https://sciwheel.com/work/citation?ids=16733256&pre=&suf=&sa=0). The OROV sequences used in this study correspond to the Genbank accession IDs: PQ168520-PQ247806 and PP153945-PQ065491. Sequence alignment for each segment (n=545) was performed using MAFFT [(*36*, *37*)](https://sciwheel.com/work/citation?ids=326539,387873&pre=&pre=&suf=&suf=&sa=0,0) and subsequently curated manually to remove artefacts using AliView [(*38*)](https://sciwheel.com/work/citation?ids=807097&pre=&suf=&sa=0). Genomic regions identified by RDP5 to have likely been acquired by recombination and genomic segments identified by RDP5 to have been acquired by reassortment, were stripped from the full genome data sets by replacing these regions with gap characters (“-”) in the alignment file, thereby a yielding a free full genome alignment free of recombination and reassortment as previously described [(*10*)](https://sciwheel.com/work/citation?ids=16733256&pre=&suf=&sa=0). Sequences with recombination and reassortment signals along the majority of the genome were completely discarded (n=43 for segment S and n=1 for segment L).

**Phylogeographic reconstruction and dispersal statistics**

To model the spatiotemporal spread of OROV using spatially-explicit phylogeographic reconstruction using the continuous diffusion model implemented in the software package BEAST 1.10 [(*40*)](https://sciwheel.com/work/citation?ids=5904397&pre=&suf=&sa=0), we utilised three different data subsets containing the 2022-2024 OROV sequences from various regions of Brazil of the S (*n* = 501), M (*n* = 545), and L (*n* = 544) segments. Before conducting the phylogeographic analyses, we assessed the strength of the molecular clock signal in each data subset using the root-to-tip regression method available in TempEst v1.5.3 [(*41*)](https://sciwheel.com/work/citation?ids=1795582&pre=&suf=&sa=0). Preliminary BEAST reconstructions revealed an outgroup of sequences which diverged from the main clade ~60 years ago for each segment; these were subsequently discarded from our analyses. Temporal structure was accepted for all datasets as the correlation coefficients were all close to or above 0.5 (S: 0.4972, M: 0.635, L: 0.5123). We reconstructed the spread of OROV lineages within Brazil by using a flexible relaxed random walk diffusion model [(*42*)](https://sciwheel.com/work/citation?ids=3052376&pre=&suf=&sa=0), which accommodates branch-specific variation in dispersal rates, with a Cauchy distribution and a jitter window size of 0.01 [(*43*)](https://sciwheel.com/work/citation?ids=16861114&pre=&suf=&sa=0). The latitude and longitude coordinates of each sample were used in this analysis. MCMC analyses were run in BEAST v1.10.4, with chains of up to 1 billion iterations each, sampling every 100,000 steps in the chain. The chains were stopped when convergence was reached following the removal of burn-in states. Convergence of each run was assessed using Tracer v1.7.1, ensuring that the effective sample size (ESS) for all relevant model parameters was >200 [(*44*)](https://sciwheel.com/work/citation?ids=5185759&pre=&suf=&sa=0). Maximum clade credibility trees were summarised using TreeAnnotator after discarding burn-in samples, the number of which was also determined in Tracer. Finally, the R package “seraphim” [(*45*)](https://sciwheel.com/work/citation?ids=10778190&pre=&suf=&sa=0) was employed to extract and map the spatiotemporal information embedded in the posterior trees. We further used “seraphim” to estimate three dispersal statistics from these movement vectors for each segment: maximal wavefront distances, weighted diffusion coefficients [(*46*)](https://sciwheel.com/work/citation?ids=2782381&pre=&suf=&sa=0), measuring the dispersal capacity of viral lineages, and an isolation-by-distance (IBD) signal measured as the Pearson correlation between the patristic and log-transformed geographic distances computed for each pair of tip nodes [(*20*)](https://sciwheel.com/work/citation?ids=16872261&pre=&suf=&sa=0).

**Ecological Niche Modelling**

Ecological niche models (ENMs) are built using a variety of statistical methods, each varying in complexity and underlying assumptions about the interaction between species occurrences and environmental factors [(*47*)](https://sciwheel.com/work/citation?ids=244931&pre=&suf=&sa=0). Recent studies have shown that disparities among different model structures can be very large, making model selection difficult [(*48*)](https://sciwheel.com/work/citation?ids=7124412&pre=&suf=&sa=0). An alternative is to use an ensemble of models to avoid selecting one single best model but instead to use a group of methods for inference. In other words, the presence of a species might be well classified by some models and misclassified by others, such that making use of an ensemble model can reduce the predictive uncertainty of a single model by combining predictions [(*49*)](https://sciwheel.com/work/citation?ids=2726135&pre=&suf=&sa=0). In this study, we applied this approach to model the distribution of OROV transmission by creating a suitability map based on the occurrence of OROV disease in Brazil and relevant environmental variables (see Geospatial Data section). For this analysis, we aggregate all raster maps to the lowest resolution available (~27km^2^). We used an ensemble of seven statistical, machine learning, and envelope models: Generalised Linear Model (GLM), Generalised Additive Model (GAM), Boosted Regression Trees (BRT), Random Forest (RF), Classification Tree Analysis (CTA), Surface Range Envelope (SRE) and Maximum Entropy Model (MAXENT). To assess the potential expansion of OROV's ecological niche in Brazil during late 2023 and 2024, we computed and compared ecological niche models (ENMs) using occurrence data from different time periods. Specifically, we created three ensemble models using pre-mid-2023 (89 occurrences), pre-2024 (133 occurrences) ,and all available occurrence records (450 occurrences).

**Supplementary Figures**

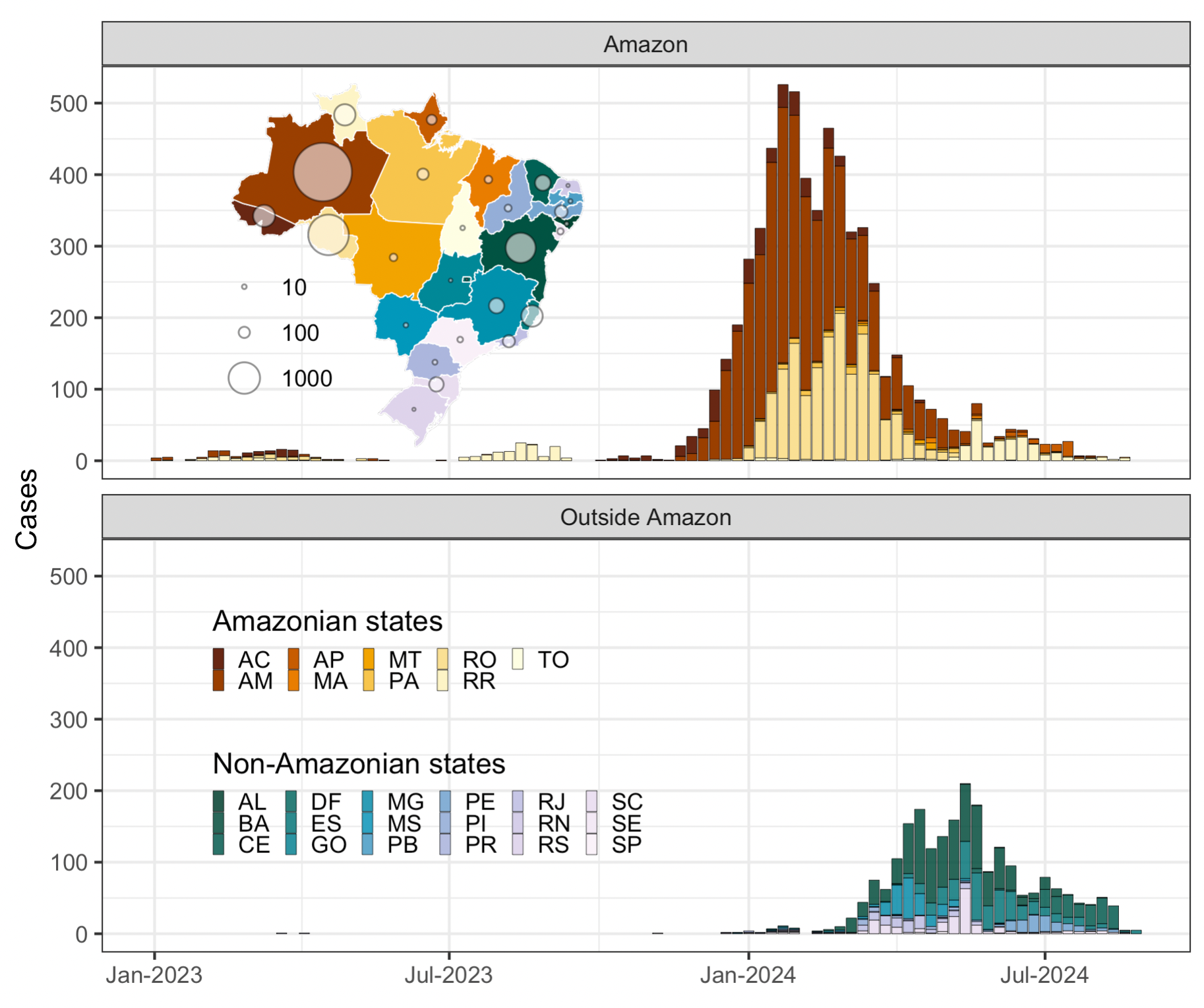

**Figure S1. Epidemiological curve of OROV cases in Brazil.** Weekly cases are shown for 2023 and 2024 divided into two epidemiological curves, one for states in the Amazon region, and one for states outside the Amazon region. The inset map is coloured by the specific state, and the circles represent the total number of recorded OROV cases in that state.

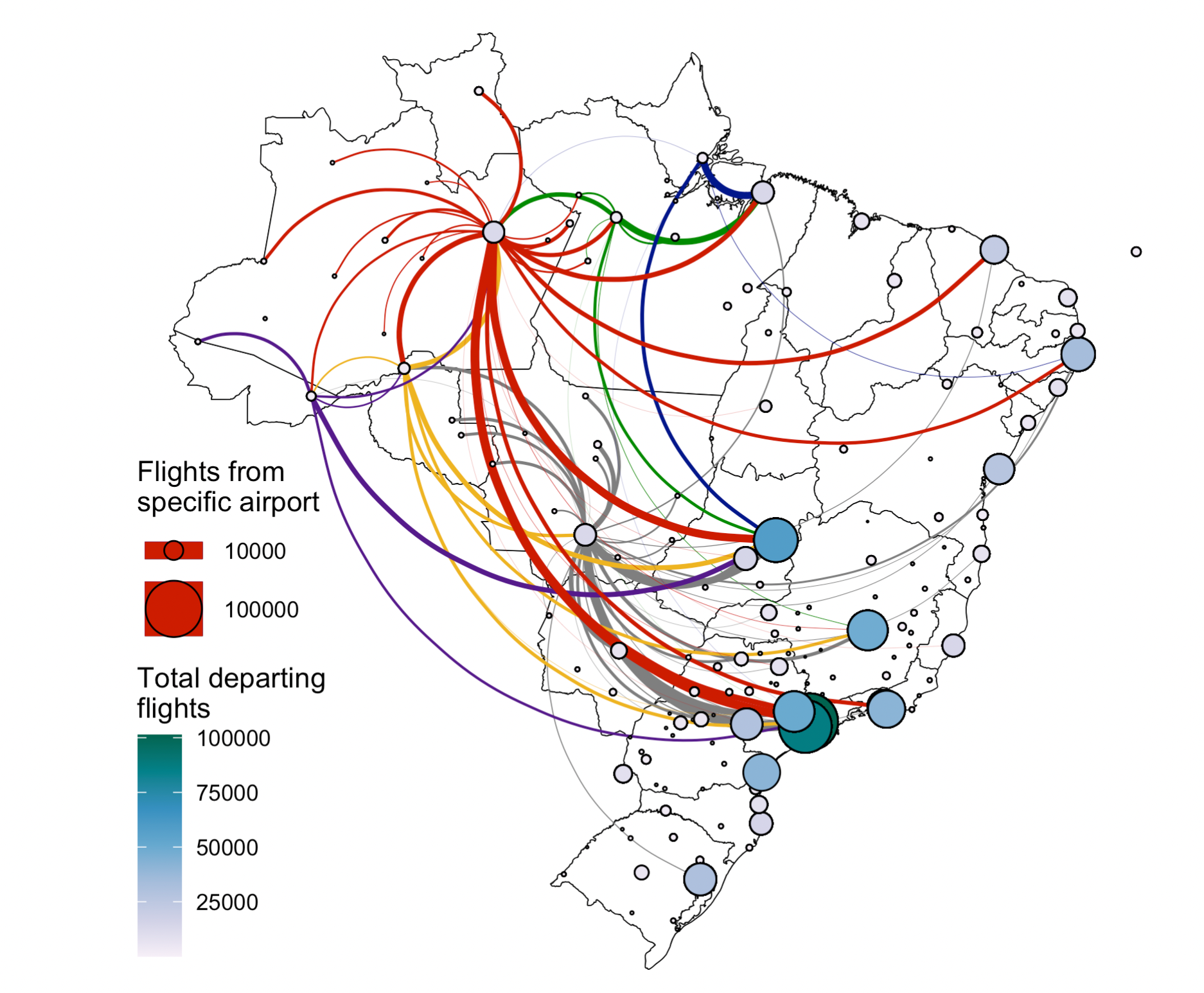

**Figure S2. Human mobility through air travel in Brazil.** The figure captures air travel data in Brazil in 2019. The map shows the total number of departing flights from all airports in Brazil. Circles are both coloured and sized by the number of flights departing from an airport origin location. The coloured curves show the number and network of flights from airports of specific municipalities, namely Manaus in state of Amazonas (red), Santarém in the state of Pará (green), Cuiabá in the state of Mato Grosso (grey), Porto Velho in the state of Rondônia (yellow), Rio Branco in the state of Acre (purple), and Macapá in the state of Amapá (blue).

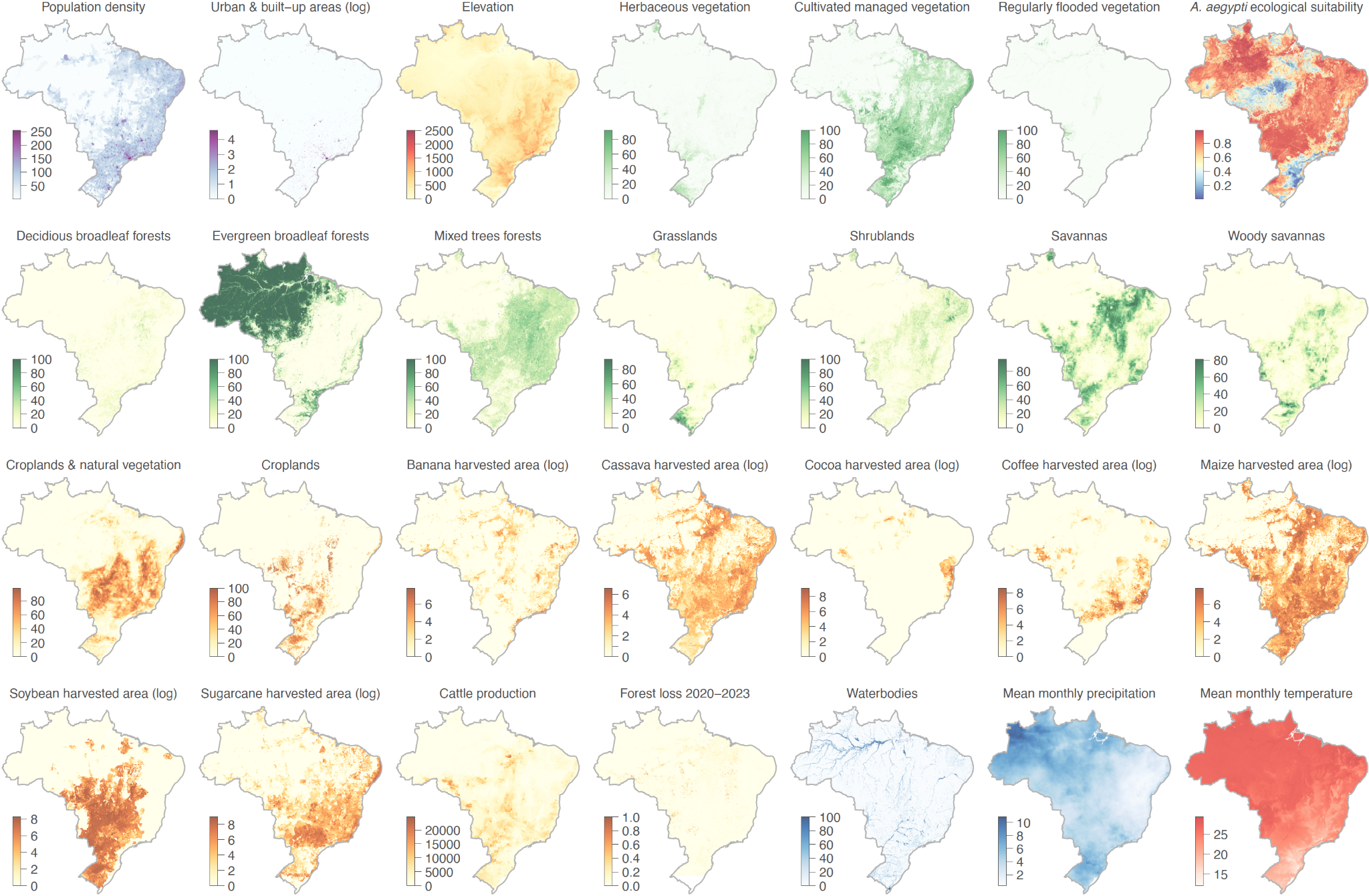
**Figure S3**. **Environmental covariates analyzed in the context of the range expansion of OROV in Brazil.** Various environmental rasters, such as demographic, land-use, and climatic covariates, were analyzed in the study to investigate their association with the spread of the OROV in Brazil. Demographic variables encompass population density and urban areas, while land-use patterns focus on the presence of croplands, water bodies, and regions impacted by deforestation linked to specific agricultural activities, such as cocoa, soy, and banana crops.

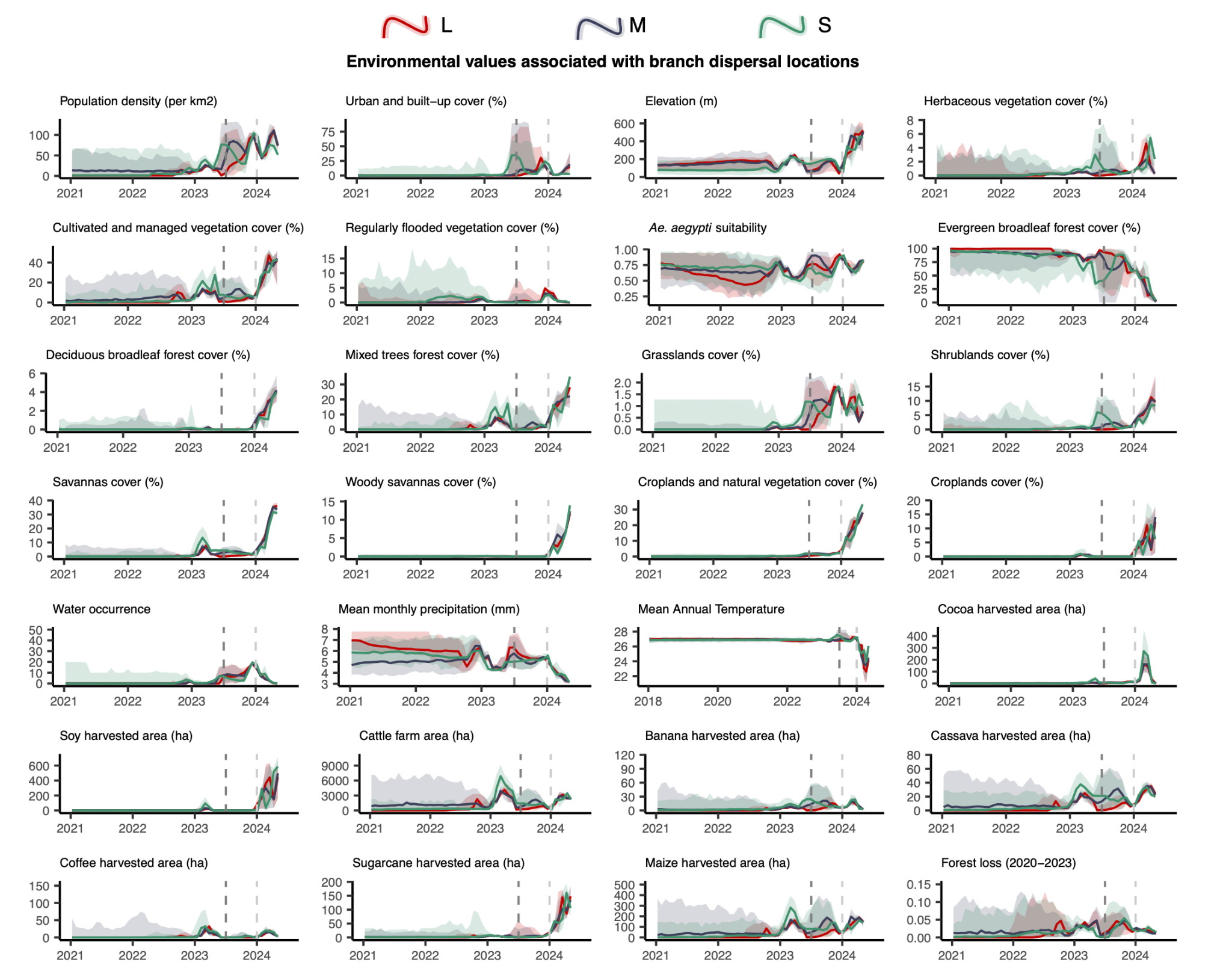
**Figure S4**. **Environmental values associated with OROV branch dispersal locations over time.** Line graphs depicting the environmental covariates associated with the locations of OROV lineage dispersal events in Brazil. Each plot illustrates how specific ecological conditions have changed over time (2021-2024) at the sites of viral lineage dispersal. This is shown for segment L (in red), segment M (in blue), and segment S (in green).

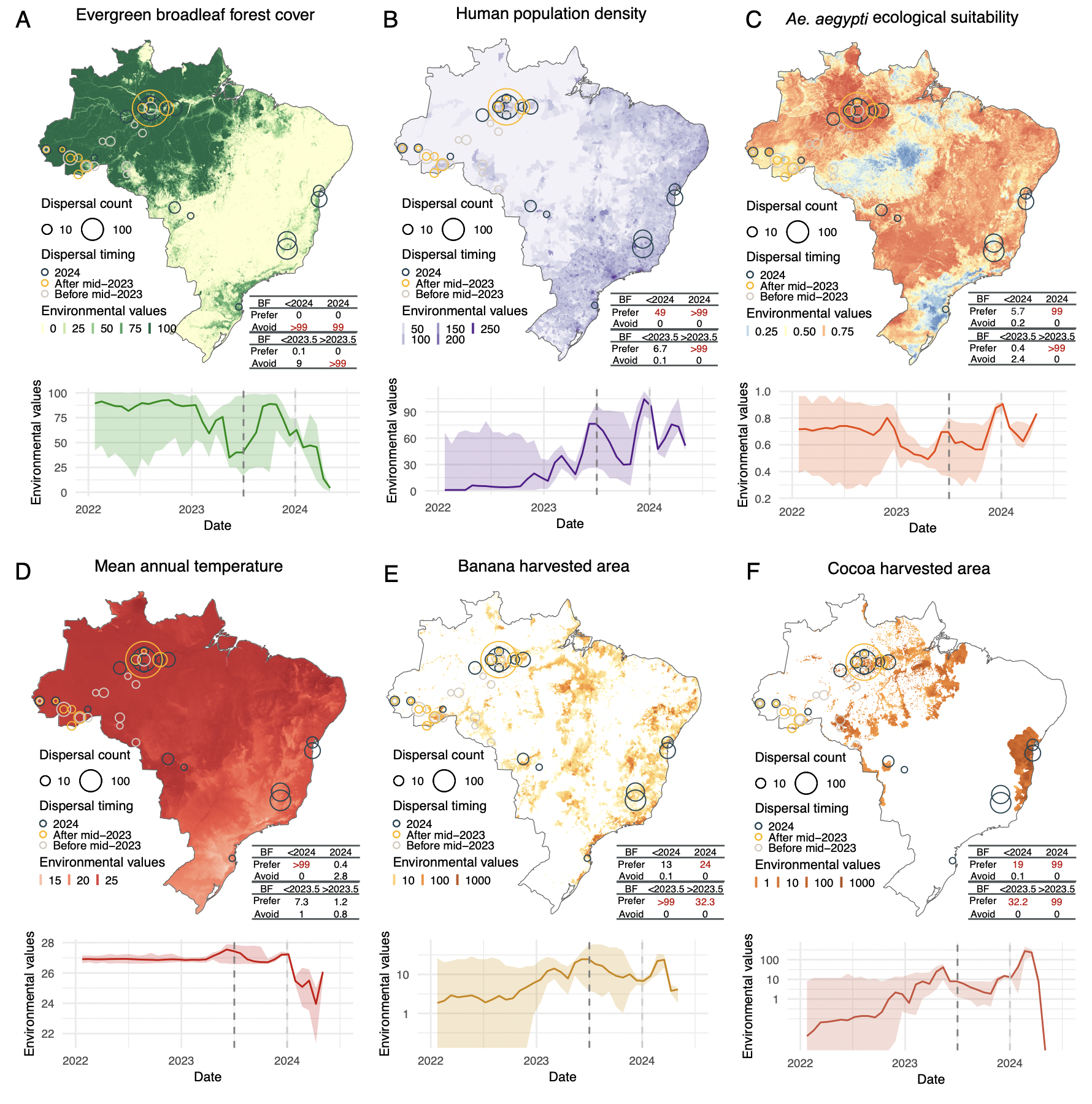

**Figure S5**. **Environmental conditions associated with OROV lineage dispersal locations over time (for segment S).** Figure panels show the spatial distribution of six main environmental factors (units specified): evergreen broadleaf forest cover (%) **(A)**, human population density (normalised between 0 and 255 per km^2^ for visual clarity) (**B**), *Ae. aegypti* ecological suitability (probability of occurrence) (**C**), mean annual temperature (°C) (**D**), banana harvested area (hectares - log) (**E**), and cocoa harvested area (hectares - log) (**F**) in the top rows. Circles on the map depict the end node of dispersal locations inferred by continuous phylogeography, sized by the number of dispersal events in an area, and coloured by the timing of the event. Bottom rows of each figure panel are line graphs depicting the environmental covariates associated with the locations of OROV lineage dispersal events in Brazil. Each plot illustrates how specific ecological conditions have changed over time (2022-2024) at the sites of viral lineage dispersal. The embedded tables show the association between environmental conditions and the dispersal location of inferred OROV lineages. Based on the analysis of 100 posterior trees obtained from continuous phylogeographic inference, the table reports Bayes factor (BF) supports for association between environmental raster values and tree node locations. Following the scale of interpretation of Kass and Raftery [(*24*)](https://sciwheel.com/work/citation?ids=948471&pre=&suf=&sa=0), we highlight BF values >20 considered as strong supports.

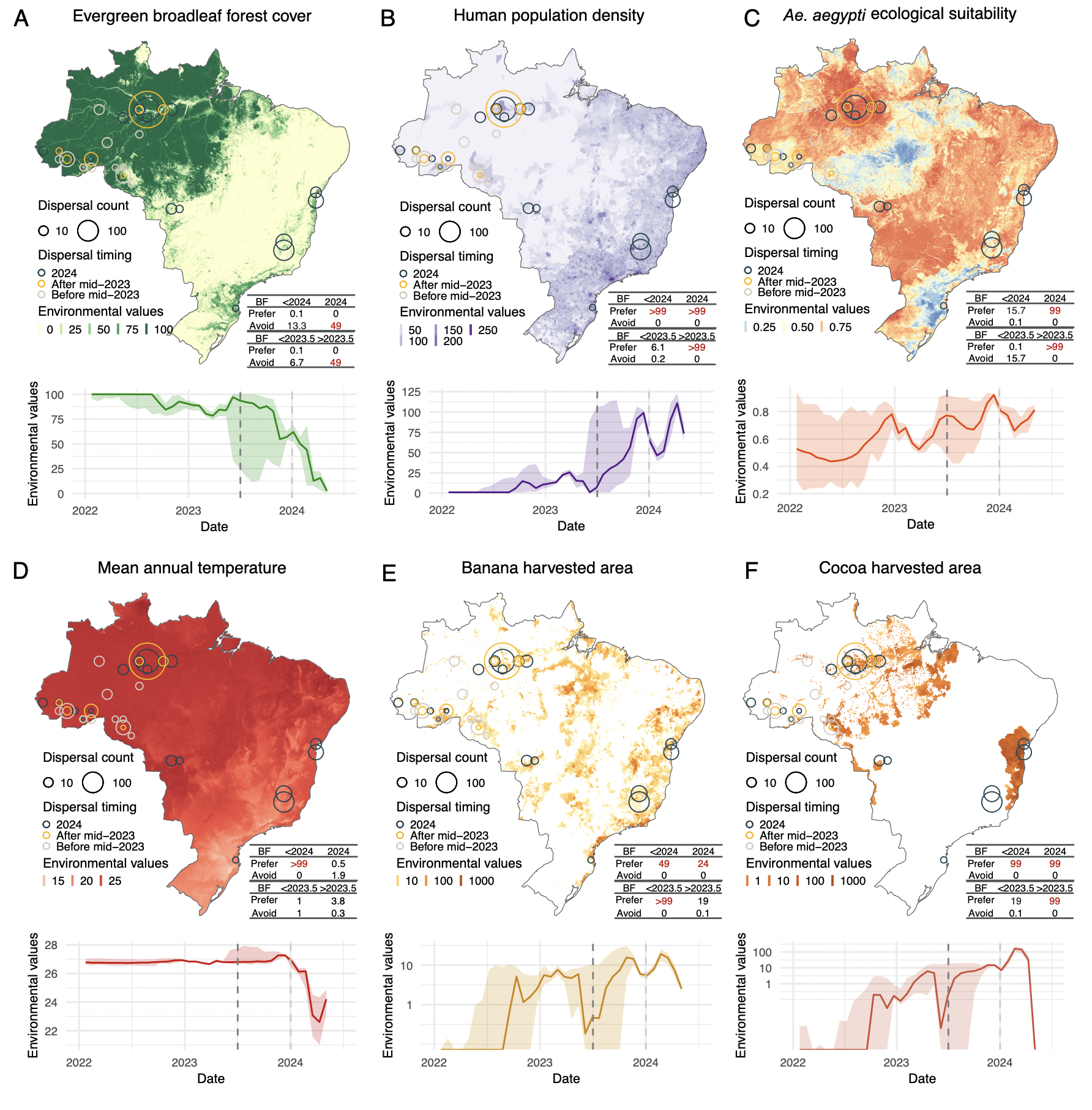

**Figure S6**. **Environmental conditions associated with OROV lineage dispersal locations over time (for segment L).** Figure panels show the spatial distribution of six main environmental factors (units specified): evergreen broadleaf forest cover (%) **(A)**, human population density (normalised between 0 and 255 per km^2^ for visual clarity) (**B**), *Ae. aegypti* ecological suitability (probability of occurrence) (**C**), mean annual temperature (°C) (**D**), banana harvested area (hectares - log) (**E**), and cocoa harvested area (hectares - log) (**F**) in the top rows. Circles on the map depict the end node of dispersal locations inferred by continuous phylogeography, sized by the number of dispersal events in an area, and coloured by the timing of the event. Bottom rows of each figure panel are line graphs depicting the environmental covariates associated with the locations of OROV lineage dispersal events in Brazil. Each plot illustrates how specific ecological conditions have changed over time (2022-2024) at the sites of viral lineage dispersal. The embedded tables show the association between environmental conditions and the dispersal location of inferred OROV lineages. Based on the analysis of 100 posterior trees obtained from continuous phylogeographic inference, the table reports Bayes factor (BF) supports for association between environmental raster values and tree node locations. Following the scale of interpretation of Kass and Raftery [(*24*)](https://sciwheel.com/work/citation?ids=948471&pre=&suf=&sa=0), we highlight BF values >20 considered as strong supports.

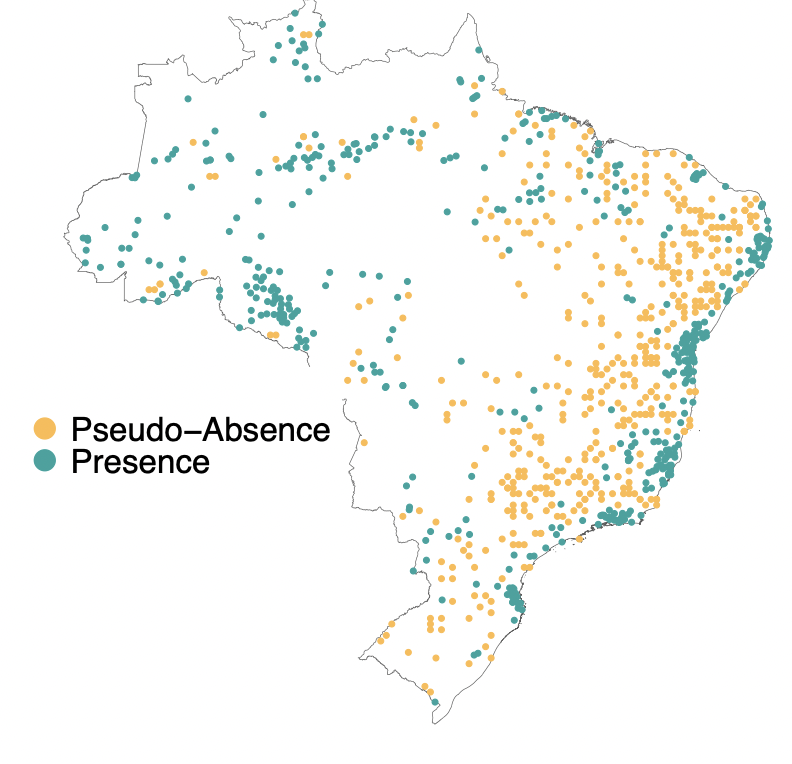

**Figure S7. Distribution of disease presence and pseudo-absence points.** We generated pseudo-absence points at a 1:1 ratio with presence points by sampling from the distribution of presence points and the kernel density estimate of human population density.

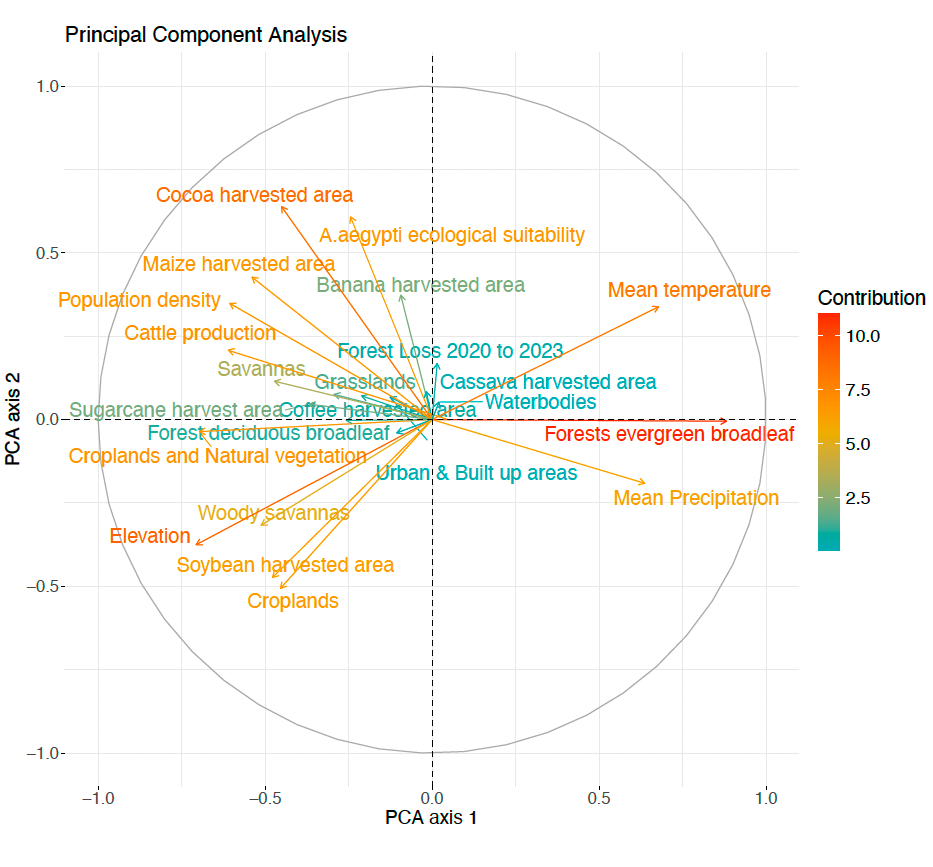

**Figure S8.** **Principal Component Analysis (PCA) plot illustrating the relationships between variables.** Arrows that lie within the same quadrant or are positioned close to each other indicate a higher correlation among the corresponding variables. Furthermore, longer arrows signify a greater contribution of those variables to the principal components, highlighting their discrimination in the overall dataset.

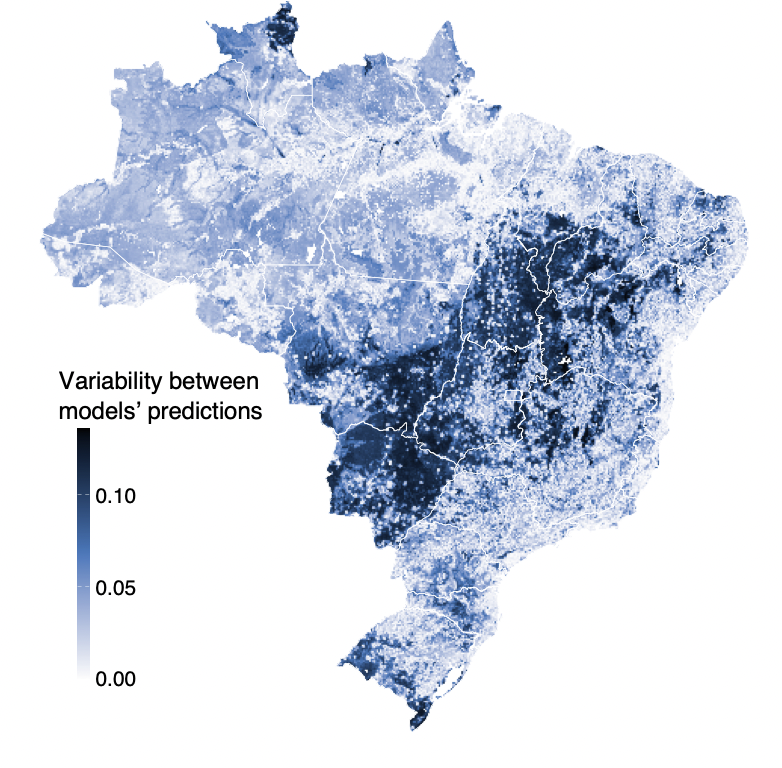

**Figure S9. Ecological niche models variability.** The degree of variability in suitability prediction values among the models in our ensemble, highlighting areas where different models either converge or diverge in their predictions.

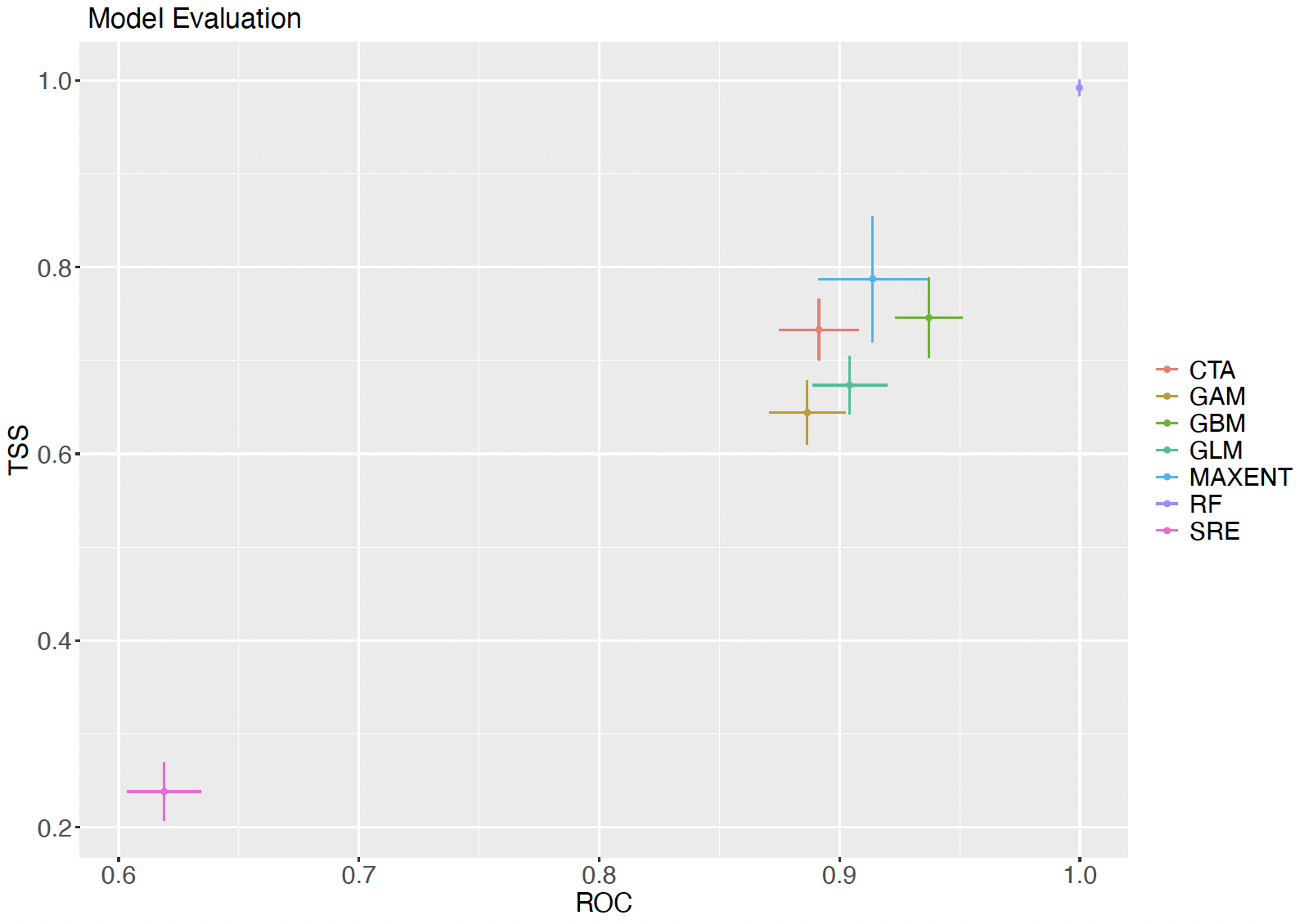

**Figure S10**. **Ecological niche models evaluation results.** Results from block cross-validation of the individual environmental niche models of the full model (using all data points). The x- and y-axis show the True Skill Statistic (TSS) and area under the Receiver Operating Characteristic (ROC) curve, respectively.

**Supplementary Table S1:**

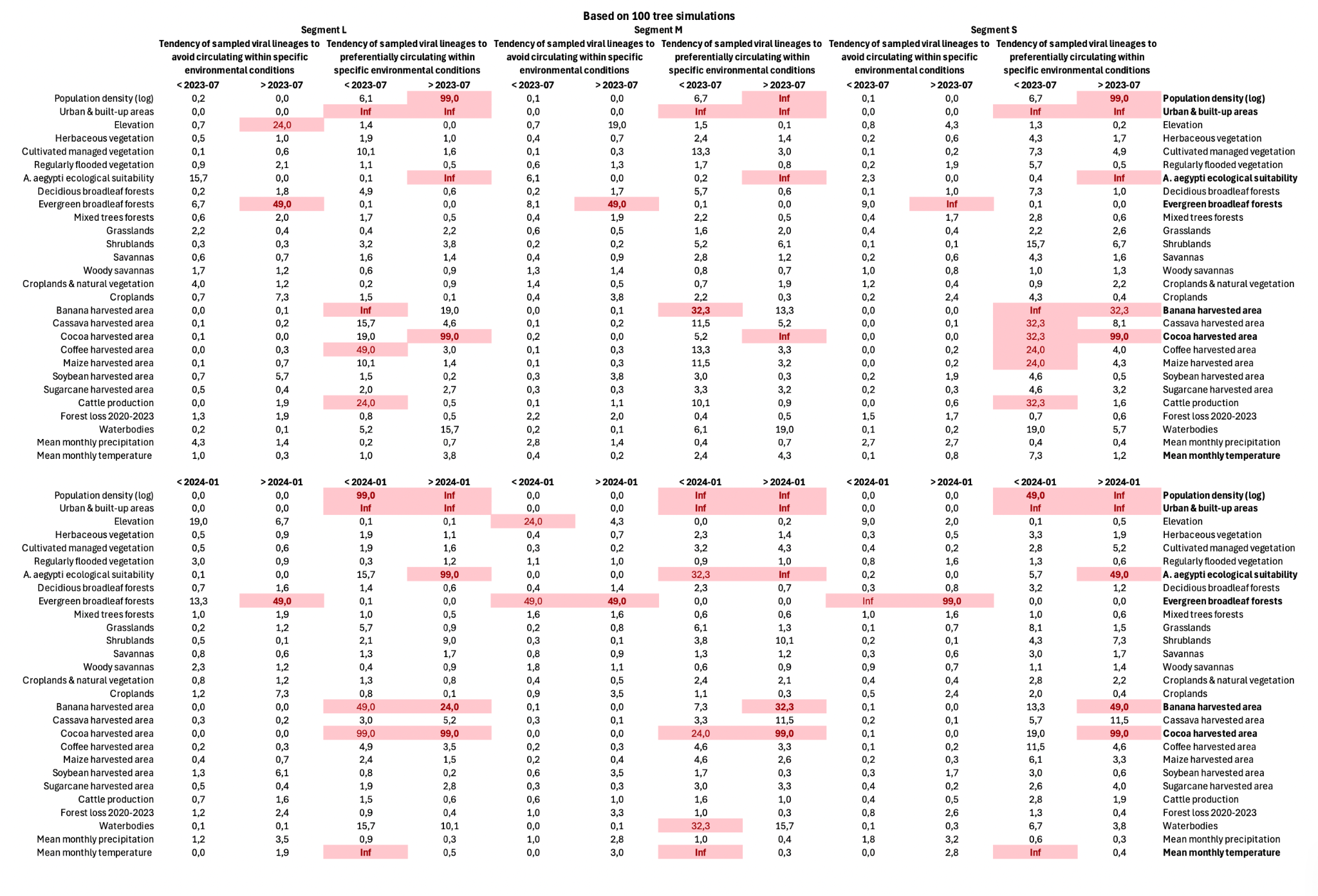

**Supplementary Table S2:** Model evaluation metrics (TSS and ROC) for the ensemble models, with results from testing on both the training/calibration datasets (left) and an independent dataset from 2024 cases (right). The models were evaluated in three stages: points collected before mid-2023, points collected before 2024, and all available points combined.

| Model performance | Occurrence Points | Testing on calibration dataset | | Testing on independent data (Year = 2024 cases) | |
| --- | --- | --- | --- | --- | --- |
| **Models** |  | **TSS** | **AUC** | **TSS** | **AUC** |
| Pre-mid-2023 | 89 | 0.77 | 0.885 | 0.4 | 0.55 |
| Pre-2024 | 133 | 0.801 | 0.957 | 0.6 | 0.862 |
| Full | 450 | 0.835 | 0.974 | 0.785 | 0.95 |

**Supplementary Table S3:** Environmental variables used in the study, with respective resolutions and data sources.

| **Environmental Variables** | **Resolution (degrees ~ km)** | **Source** |
| --- | --- | --- |
| Population density | 0.0083 ~ 1 km | WorldPop, Global Human Settlement Layer (GHSL) |
| Urban & built up areas cover | 0.0083 ~ 1 km | GHSL, World Urbanization Prospects |
| Annual mean temperature | 0.0090 ~ 1 km | WorldClim |
| Elevation | 0.0083 ~ 1 km | USGS |
| Herbaceous vegetation cover | 0.0083 ~ 1 km | MODIS Land Cover, Global Land Cover (GLC) |
| Cultivated managed vegetation cover | 0.0083 ~ 1 km | EarthEnv - https://www.earthenv.org/landcover |
| Regularly flooded vegetation cover | 0.0083 ~ 1 km | EarthEnv - https://www.earthenv.org/landcover |
| *Ae. aegypti* ecological suitability | 0.0146 ~ 1.62 km | DOI: https://doi.org/10.7554/eLife.08347 |
| Deciduous broadleaf forest cover | 0.0083 ~ 1 km | EarthEnv - https://www.earthenv.org/landcover |
| Evergreen broadleaf forest cover | 0.0083 ~ 1 km | EarthEnv - https://www.earthenv.org/landcover |
| Mixed trees forest cover | 0.0083 ~ 1 km | EarthEnv - https://www.earthenv.org/landcover |
| Grasslands cover | 0.25 ~ 27.75 km | EarthEnv - https://www.earthenv.org/landcover |
| Shrublands cover | 0.0083 ~ 1 km | EarthEnv - https://www.earthenv.org/landcover |
| Savannas cover | 0.25 ~ 27.75 km | EarthEnv - https://www.earthenv.org/landcover |
| Woody savannas cover | 0.25 ~ 27.75 km | EarthEnv - https://www.earthenv.org/landcover |
| Croplands & natural vegetation cover | 0.25 ~ 27.75 km | EarthEnv - https://www.earthenv.org/landcover |
| Croplands cover | 0.25 ~ 27.75 km | EarthEnv - https://www.earthenv.org/landcover |
| Banana harvested area | 0.0833 ~10 km | EarthEnv - https://www.earthenv.org/landcover |
| Cassava harvested area | 0.0833 ~10 km | EarthEnv - https://www.earthenv.org/landcover |
| Cocoa harvested area | 0.0833 ~10 km | EarthEnv - https://www.earthenv.org/landcover |
| Coffee harvested area | 0.0833 ~10 km | EarthEnv - https://www.earthenv.org/landcover |
| Maize harvested area | 0.0833 ~10 km | EarthEnv - https://www.earthenv.org/landcover |
| Soybean harvested area | 0.0833 ~10 km | EarthEnv - https://www.earthenv.org/landcover |
| Sugarcane harvested area | 0.0833 ~10 km | EarthEnv - https://www.earthenv.org/landcover |
| Cattle cultivation area | 0.0833 ~10 km | EarthEnv - https://www.earthenv.org/landcover |
| Forest loss (2020-2023) | 0.025 ~ 2.78 km | https://glad.earthengine.app/view/global-forest-change |
| Water occurrence | 0.0833 ~ 10 km | Global surface water explorer |
| Annual mean precipitation | 0.05 ~ 5.55 km | WorldClim |

**Supplementary References**

[36.	K. Katoh, K. Misawa, K. Kuma, T. Miyata, MAFFT: A novel method for rapid multiple sequence alignment based on fast Fourier transform. *Nucleic Acids Res.* **30**, 3059–3066 (2002).](https://sciwheel.com/work/bibliography/326539)

[37.	K. Katoh, D. M. Standley, MAFFT multiple sequence alignment software version 7: improvements in performance and usability. *Mol. Biol. Evol.* **30**, 772–780 (2013).](https://sciwheel.com/work/bibliography/387873)

[38.	A. Larsson, AliView: a fast and lightweight alignment viewer and editor for large datasets. *Bioinformatics*. **30**, 3276–3278 (2014).](https://sciwheel.com/work/bibliography/807097)

[39.	Brazil’s Air Travel Hits Pre-Pandemic Heights - The Rio Times, (available at https://www.riotimesonline.com/brazil-news/brazils-air-travel-hits-pre-pandemic-heights/).](https://sciwheel.com/work/bibliography/17100260)

[40.	M. A. Suchard, P. Lemey, G. Baele, D. L. Ayres, A. J. Drummond, A. Rambaut, Bayesian phylogenetic and phylodynamic data integration using BEAST 1.10. *Virus Evol.* **4**, vey016 (2018).](https://sciwheel.com/work/bibliography/5904397)

[41.	A. Rambaut, T. T. Lam, L. Max Carvalho, O. G. Pybus, Exploring the temporal structure of heterochronous sequences using TempEst (formerly Path-O-Gen). *Virus Evol.* **2**, vew007 (2016).](https://sciwheel.com/work/bibliography/1795582)

[42.	P. Lemey, A. Rambaut, J. J. Welch, M. A. Suchard, Phylogeography takes a relaxed random walk in continuous space and time. *Mol. Biol. Evol.* **27**, 1877–1885 (2010).](https://sciwheel.com/work/bibliography/3052376)

[43.	S. Dellicour, M. S. Gill, N. R. Faria, A. Rambaut, O. G. Pybus, M. A. Suchard, P. Lemey, Relax, Keep Walking - A Practical Guide to Continuous Phylogeographic Inference with BEAST. *Mol. Biol. Evol.* **38**, 3486–3493 (2021).](https://sciwheel.com/work/bibliography/16861114)

[44.	A. Rambaut, A. J. Drummond, D. Xie, G. Baele, M. A. Suchard, Posterior summarization in Bayesian phylogenetics using Tracer 1.7. *Syst. Biol.* **67**, 901–904 (2018).](https://sciwheel.com/work/bibliography/5185759)

[45.	S. Dellicour, R. Rose, N. R. Faria, P. Lemey, O. G. Pybus, SERAPHIM: studying environmental rasters and phylogenetically informed movements. *Bioinformatics*. **32**, 3204–3206 (2016).](https://sciwheel.com/work/bibliography/10778190)

[46.	N. S. Trovão, M. A. Suchard, G. Baele, M. Gilbert, P. Lemey, Bayesian Inference Reveals Host-Specific Contributions to the Epidemic Expansion of Influenza A H5N1. *Mol. Biol. Evol.* **32**, 3264–3275 (2015).](https://sciwheel.com/work/bibliography/2782381)

[47.	C. Merow, M. J. Smith, J. A. Silander, A practical guide to MaxEnt for modeling species’ distributions: what it does, and why inputs and settings matter. *Ecography*. **36**, 1058–1069 (2013).](https://sciwheel.com/work/bibliography/244931)

[48.	A. Guisan, W. Thuiller, N. E. Zimmermann, *Habitat suitability and distribution models: with applications in R* (Cambridge University Press, Cambridge, 2017), *Ecology, Biodiversity and Conservation*.](https://sciwheel.com/work/bibliography/7124412)

[49.	M. Marmion, M. Parviainen, M. Luoto, R. K. Heikkinen, W. Thuiller, Evaluation of consensus methods in predictive species distribution modelling. *Diversity and Distributions*. **15**, 59–69 (2009).](https://sciwheel.com/work/bibliography/2726135)

[50.	R. Muscarella, P. J. Galante, M. Soley-Guardia, R. A. Boria, J. M. Kass, M. Uriarte, R. P. Anderson, ENMeval: An R package for conducting spatially independent evaluations and estimating optimal model complexity for Maxent ecological niche models. *Methods Ecol. Evol.* **5**, 1198–1205 (2014).](https://sciwheel.com/work/bibliography/2459809)

[51.	B. Gregorutti, B. Michel, P. Saint-Pierre, Correlation and variable importance in random forests. *Stat. Comput.* **27**, 659–678 (2017).](https://sciwheel.com/work/bibliography/13573266)

**CLIMADE Consortium Contributing Authors:**
